# Transition to Endemic: Acceptance of Additional COVID-19 Vaccine Doses Among Canadian Adults in A National Cross-Sectional Survey

**DOI:** 10.1101/2022.06.27.22276870

**Authors:** Laura Reifferscheid, Janet Sau Wun Lee, Noni E MacDonald, Manish Sadarangani, Ali Assi, Samuel Lemaire-Paquette, Shannon E. MacDonald

## Abstract

**Background:** Additional doses of COVID-19 vaccine have been proposed as solutions to waning immunity and decreased effectiveness of primary doses against infection with new SARS-CoV-2 variants. However, the effectiveness of additional vaccine doses relies on widespread population acceptance. We aimed to assess the acceptance of additional COVID-19 vaccine doses (third and annual doses) among Canadian adults and determine associated factors.

**Methods:** We conducted a national, cross-sectional online survey among Canadian adults from October 14 to November 12, 2021. Weighted multinomial logistic regression analyses were used to identify sociodemographic and health-related factors associated with third and annual dose acceptance and indecision, compared to refusal. We also assessed influences on vaccine decision-making, and preferences for future vaccine delivery.

**Results:** Of 6010 respondents, 70% reported they would accept a third dose, while 15.2% were undecided. For annual doses, 64% reported acceptance, while 17.5% were undecided. Factors associated with third dose acceptance and indecision were similar to those associated with annual dose acceptance and indecision. Previous COVID-19 vaccine receipt, no history of COVID-19 disease, intention to receive an influenza vaccine, and increasing age were strongly associated with both acceptance and indecision. Chronic illness was associated with higher odds of acceptance, while self-reported disability was associated with higher odds of being undecided. Higher education attainment and higher income were associated with higher odds of accepting additional doses. Minority first language was associated with being undecided about additional doses, while visible minority identity was associated with being undecided about a third dose and refusing an annual dose. All respondents reported government recommendations were an important influence on their decision-making and identified pharmacy-based delivery and drop-in appointments as desirable. Co-administration of COVID-19 and influenza vaccines was viewed positively by 75.5% of the dose 3 acceptance group, 12.3% of the undecided group, and 8.4% of the refusal group.

**Conclusions:** To increase acceptance, targeted interventions among visible minority and minority language populations, and those with a disability, are required. Offering vaccination at pharmacies and through drop-in appointments are important to facilitate uptake, while offering COVID-19/influenza vaccine co-administration may have little benefit among those undecided about additional doses.

## Background

COVID-19 vaccines have proven to be highly effective at limiting morbidity and mortality associated with SARS-CoV-2 infection.(1) However, the ongoing emergence of new SARS-CoV-2 variants of concern, and waning immunity among already vaccinated individuals, have highlighted the need for additional vaccine doses to boost population immunity.(2-4) In settings where primary series coverage exceeds 50%, booster dose programs may be a more effective strategy for limiting population-wide negative effects of COVID-19 disease than improving primary series uptake among the unvaccinated.(5) This has led some countries, including Canada (6-7), to consider additional COVID-19 vaccine doses as a key strategy to combat the evolving COVID-19 pandemic.

While the scientific evidence for boosters is encouraging, this strategy requires population acceptance and significant public health efforts. Even among those who have already received a two-dose series, there is a considerable portion of the population who are unwilling or undecided about receiving a third dose.(8-11) However, adult perceptions on ongoing booster doses (i.e., annual COVID-19 vaccination) are largely unknown, and evidence on additional vaccine dose acceptance among Canadian adults, particularly among populations who may be at greater risk for COVID-19 infection and/or severe illness, is required. To ensure that COVID-19 vaccination programs are both acceptable and sustainable, we need an understanding of perceptions on strategies used to promote vaccine uptake over the long term, including vaccination mandates and restrictions, vaccination locations, and COVID-19 vaccine co-administration with other vaccines.(12)

The aim of this study was to determine Canadian adults’ intention for receiving additional doses of COVID-19 vaccine (i.e., third or annual doses), and examine factors associated with acceptance. We further sought to understand influences on vaccination decision-making, and preferences around vaccine delivery location, and COVID-19 vaccine co-administration.

## Methods

### Study Design and Setting

We conducted an online, cross-sectional survey among Canadian adults from October 14 to November 12, 2021. At the time of this study, Canada was experiencing the “fourth wave” of the COVID-19 pandemic, driven largely by the Delta variant(13), and concerns around the Omicron variant were starting to develop.(14) Jurisdictions across Canada were focusing on improving vaccine uptake, with many requiring proof of vaccination for certain groups to access certain businesses, venues, and activities.(13) Approximately 87% of adults had received at least one dose of COVID-19 vaccine (15), and a third dose had only been recommended for immunocompromised individuals and other select groups, based on level of disease risk.(16) Vaccines requiring only a one dose primary series were not approved for use in Canada at that time.(17)

### Sample

Respondents were drawn from a national polling panel of >400,000 Canadians (18), recruited to closely match the proportion of the Canadian population by region of residence, age, and sex.(19) We purposefully recruited minimum quotas of target populations of interest, in order to include populations that were prioritized for additional doses of COVID-19 vaccine, have experienced disproportionately high rates of COVID-19 infection, and those often underrepresented in research. Target populations included: parents/caregivers (defined as having one or more children aged 0-17 in their home), Indigenous persons, visible minorities, those with a minority first language, newcomers (defined as arriving to Canada within the past 5 years), persons with chronic medical conditions, persons with disabilities, and healthcare workers. Based on the maximum variability possible in the outcome variable in the population (i.e., a proportion of 0.50), with a margin of error of +/-5% and 95% confidence intervals (CI), the minimum sample size for each target population was estimated to be 402 respondents. In order to complete the survey, internet access and the ability to read either English or French was required. All respondents aged 18 years and older were included in this analysis.

### Data Collection

Online survey questions were developed based on a previous national survey about perceptions and intentions for COVID-19 vaccination (20), areas of focus for our policy partners, and the expertise of our national team of immunization researchers and policy advisors. The survey was reviewed by public health experts for content validity and tested for readability and usability by team members. The survey was also pilot tested with 47 members of the public; revisions were made based on their feedback. Survey questions used in this study are provided in Table A1.

Quality control efforts to promote rigour and validity of survey responses included creation of a unique URL identifier for each respondent, telephone follow-up for identity confirmation with 15% of the respondents, and embedded consistency questions to identify and eliminate inattentive respondents.(18) Ethics approval for this study was received from the Health Research Ethics Board at the University of Alberta.

### Measures

All respondents were asked about their current COVID-19 vaccine status, defined as having received none, one, or two doses of COVID-19 vaccine. Respondents who had received at least one dose were then asked about their intention to receive additional doses. Our outcome variable of “COVID-19 third dose intention” was coded as acceptance (those who responded “yes”), undecided (those who were “undecided”), and rejection (those who responded “no” and those who had not yet received any doses of COVID-19 vaccine). Using the same method, we constructed an outcome variable for “COVID-19 annual dose intention”.

We examined the association of our two outcome variables with a number of exposure variables. Sociodemographic variables included age, gender, region of residence, self-reported race and ethnicity, level of education, and annual household income. Respondents were also asked about the first language they learned to speak (to differentiate between French/English [Canada’s official languages] and a minority language), parent/caregiver status, length of time in Canada, and if they were employed in healthcare. Participants were also asked to indicate any disability, chronic condition, previous COVID-19 disease, and COVID-19 vaccination status.

All respondents were asked to identify important influences on their COVID-19 vaccination decision. Respondents who had received at least one dose of COVID-19 vaccine were further asked to identify their main reason for having received a COVID-19 vaccine, their perceptions of COVID-19 vaccine co-administration, and ways to make future vaccinations easier. Where collected, free-text responses were coded into existing categories or new categories.

### Statistical Analysis

Survey results were weighted to more accurately represent Canada’s national population, following Leger’s standard protocol. Raking, an iterative proportional fitting method, was used to calculate weights based on age, gender and province of residence according to data from the 2016 Canadian Census.(19) As we had included targeted sampling of particular population groups in our sampling method, weights were adjusted to ensure our overall sample did not overrepresent any of these groups. For these target variables, weighting was based on Census data when available (i.e., race/ethnicity, newcomer status, and first language). For variables without corresponding Census data, weighting estimates were based on standard processes established by the survey company, drawing on total survey attempts and results from weekly panel-wide surveys.(18)

We used unweighted and weighted data to calculate descriptive statistics for all variables, including frequencies and percentages for categorical variables and means and standard deviations for continuous variables. Using weighted data, we developed bivariate and multivariate multinomial logistic regression models to determine the factors associated with each of our two outcome variables (COVID-19 dose 3 intention, COVID-19 annual dose intention), presented as adjusted odds ratios (aOR) and 95% confidence intervals (CI). For both outcome variables, we compared those who intended to receive the vaccine (acceptance group) and those who were undecided (undecided group), to those who did not intend to receive the vaccine (refusal group). The continuous age variable met the assumption of linearity in the final model. Region of residence was removed from the final model as the AIC was significantly higher with it included. All included variables were confirmed to provide unique information (defined as variance inflation factor <5). ‘Prefer not to answer’ responses were excluded from the regression analyses via listwise deletion. Descriptive analyses were completed using SPSS version 26.0 (IBM, Chicago, IL, USA) and regressions were completed using R version 4.0.2 (R Foundation, Vienna, AT).

## Results

The survey was completed by 6010 adult respondents. Unweighted and weighted characteristics and vaccination intentions for the respondents are presented in Table 1. The majority of respondents (70.3%) reported that they planned to receive a third dose of the vaccine, while 15.2% and 14.5% reported they were undecided or refused to receive a third dose, respectively. Most respondents (64.7%) also reported acceptance of an annual COVID-19 vaccine dose, with the remaining respondents divided between undecided (17.5%) and refusal (17.8%).

**Table 1.** Respondent characteristics and COVID-19 vaccine additional dose acceptance (unweighted and weighted)

| Characteristic | Unweighted (N=6010) |  | Weighted |
| --- | --- | --- | --- |
|  | n | % | % (95% CI) |
| **Age** |  |  |  |
| 18-24 | 409 | 6.8 | 12.8 (12.0-13.7) |
| 25-34 | 1268 | 21.1 | 17.1 (16.1-18.1) |
| 35-44 | 1499 | 24.9 | 15.6 (14.7-16.5) |
| 45-54 | 1284 | 21.4 | 17.3 (16.3-19.2) |
| 55-59 | 444 | 7.4 | 9.0 (8.3-9.7) |
| 60-69 | 650 | 10.8 | 14.6 (13.7-15.5) |
| 70+ | 456 | 7.6 | 13.6 (12.7-14.5) |
| **Gender** |  |  |  |
| Woman | 3425 | 57.0 | 51.1 (49.9-52.4) |
| Man | 2545 | 42.3 | 48.0 (46.8-49.3) |
| Other<sup>a</sup> | 40 | 0.7 | 0.8 (0.6-1.1) |
| **Region of residence** |  |  |  |
| British Columbia | 743 | 12.4 | 13.6 (12.7-14.5) |
| Alberta | 651 | 10.8 | 11.2 (10.4-12.0) |
| Saskatchewan | 156 | 2.6 | 2.6 (2.2-3.0) |
| Manitoba | 235 | 3.9 | 4.0 (3.5-4.5) |
| Ontario | 2103 | 35.0 | 38.5 (37.3-39.7) |
| Quebec | 1724 | 28.7 | 23.3 (22.3-24.4) |
| Atlantic<sup>b</sup> | 398 | 6.6 | 6.8 (6.2-7.4) |
| **Self-reported race and ethnicity<sup>c</sup>** |  |  |  |
| White | 4004 | 67.7 | 73.1 (72.0-74.2) |
| Visible minority | 1408 | 23.8 | 21.9 (20.8-22.9) |
| Indigenous<sup>d</sup> | 503 | 8.5 | 5.0 (4.5-5.6) |
| Prefer not to answer | 95 | 1.6 | N/A |
| **First language** |  |  |  |
| English/French | 5279 | 87.8 | 79.0 (78.0-80.0) |
| Other | 731 | 12.2 | 21.0 (20.0-22.0) |
| **Newcomer<sup>e</sup>** |  |  |  |
| Yes | 506 | 8.4 | 3.5 (3.1-4.0) |
| No | 5504 | 91.6 | 96.5 (96.0-96.9) |
| **Parent** |  |  |  |
| Yes | 2530 | 42.1 | 30.2 (29.1-31.4) |
| No | 3480 | 57.9 | 69.8 (68.6-70.9) |
| **Disability<sup>f</sup>** |  |  |  |
| Yes | 887 | 14.8 | 17.3 (16.3-18.2) |
| No or don't know | 5097 | 84.8 | 82.7 (81.8-83.7) |
| Prefer not to answer | 26 | 0.4 | N/A |
| **Chronic illness<sup>g</sup>** |  |  |  |
| Yes | 1555 | 25.9 | 26.2 (25.1-27.3) |
| No | 4455 | 74.1 | 73.8 (72.7-74.9) |
| **Healthcare worker** |  |  |  |
| Yes | 694 | 11.5 | 8.4 (7.7-9.1) |
| No | 5316 | 88.5 | 91.6 (90.9-92.3) |
| **Annual household income** |  |  |  |
| <40,000 | 1037 | 17.3 | 20.8 (19.8-21.8) |
| 40,000-79,999 | 1610 | 26.8 | 30.7 (29.6-31.9) |
| 80,000+ | 2801 | 46.6 | 48.5 (47.2-49.7) |
| Prefer not to answer | 562 | 9.4 | N/A |
| **Education** |  |  |  |
| High school or less | 1001 | 16.7 | 19.8 (18.8-20.8) |
| Non-university certificate or diploma | 1872 | 31.1 | 29.3 (28.2-30.5) |
| University certificate, bachelor's degree, post-graduate degree | 3102 | 51.6 | 50.9 (49.6-52.2) |
| Prefer not to answer | 35 | 0.6 | N/A |
| **Plan to receive seasonal influenza vaccine** |  |  |  |
| Disagree | 2001 | 33.3 | 30.4 (29.3-31.6) |
| Neutral | 905 | 15.1 | 14.8 (13.9-15.7) |
| Agree | 3104 | 51.6 | 54.7 (53.4-56.0) |

| **COVID-19 vaccination status** |  |  |  |
| --- | --- | --- | --- |
| No doses | 519 | 8.6 | 8.6 (7.9-9.3) |
| One dose | 183 | 3.0 | 2.8 (2.3-3.2) |
| Two doses | 5308 | 88.3 | 88.6 (87.8-89.4) |
| **COVID-19 disease history** |  |  |  |
| No or Don't know | 5378 | 89.5 | 90.1 (89.3-90.8) |
| Yes or Think so, but not confirmed | 620 | 10.3 | 9.9 (9.2-10.7) |
| **COVID-19 dose 3 intentions** |  |  |  |
| No | 949 | 15.8 | 14.5 (13.6-15.4) |
| Undecided | 967 | 16.1 | 15.2 (14.3-16.1) |
| Yes | 4094 | 68.1 | 70.3 (69.1-71.5) |
| **COVID-19 annual dose intentions** |  |  |  |
| No | 1183 | 19.7 | 17.8 (16.8-18.7) |
| Undecided | 1139 | 19.0 | 17.5 (16.6-18.5) |
| Yes | 3688 | 61.4 | 64.7 (63.5-65.9) |

*CI*, confidence interval; *N/A*, not applicable
<sup>a</sup> Includes options for gender non-conforming, transgender, two-spirit, and open-ended responses
<sup>b</sup> Nova Scotia, New Brunswick, Prince Edward Island, and Newfoundland and Labrador
<sup>c</sup> Self-identified as White or visible minority groups (e.g. Black, Latin/Central American, Arabic/West Asian/North African, East Asian, South Asian, Other) as per Statistics Canada (2021). Visible minorities are defined as non-White and non-Indigenous (Statistics Canada, 2021).
<sup>d</sup> Self-identified as First Nations, Métis, or Inuk (the three Indigenous populations in Canada)
<sup>e</sup> Arrived in Canada within the past 5 years
<sup>f</sup> Self-reported limitation in type or amount of activity because of a long-term physical condition, mental condition, or health problem
<sup>g</sup> Pre-existing chronic conditions defined as severe asthma requiring medical follow-up or hospitalization, other severe chronic lung disease requiring regular medical follow-up or hospitalization (e.g. emphysema, chronic bronchitis, or cystic fibrosis), severe heart problem requiring regular medical follow-up or hospitalization (e.g. angina, heart failure, heart attack), diabetes, liver disease, chronic kidney disease, cancer or other immune system disorder, immunocompromised state from organ transplant or immune deficiencies, obesity, dementia.

### Factors associated with accepting or being undecided about third dose COVID-19 vaccination

In the multivariable model (Table 2), increasing age, positive or neutral intention to receive a seasonal influenza vaccine, previous receipt of a COVID-19 vaccine (one or two doses), and no history of COVID-19 disease were all associated with both accepting or being undecided about receiving a third dose of COVID-19 vaccine (compared to refusal). Non-parent status, presence of a chronic illness, and a higher level of educational attainment were also associated with higher odds of accepting a third dose, but not significantly associated with being undecided about third dose receipt. Identifying as a visible minority, minority (i.e., non-English or French) first language, and presence of a disability were also associated with higher odds of being undecided about receiving a third dose, compared with refusal.

**Table 2.**
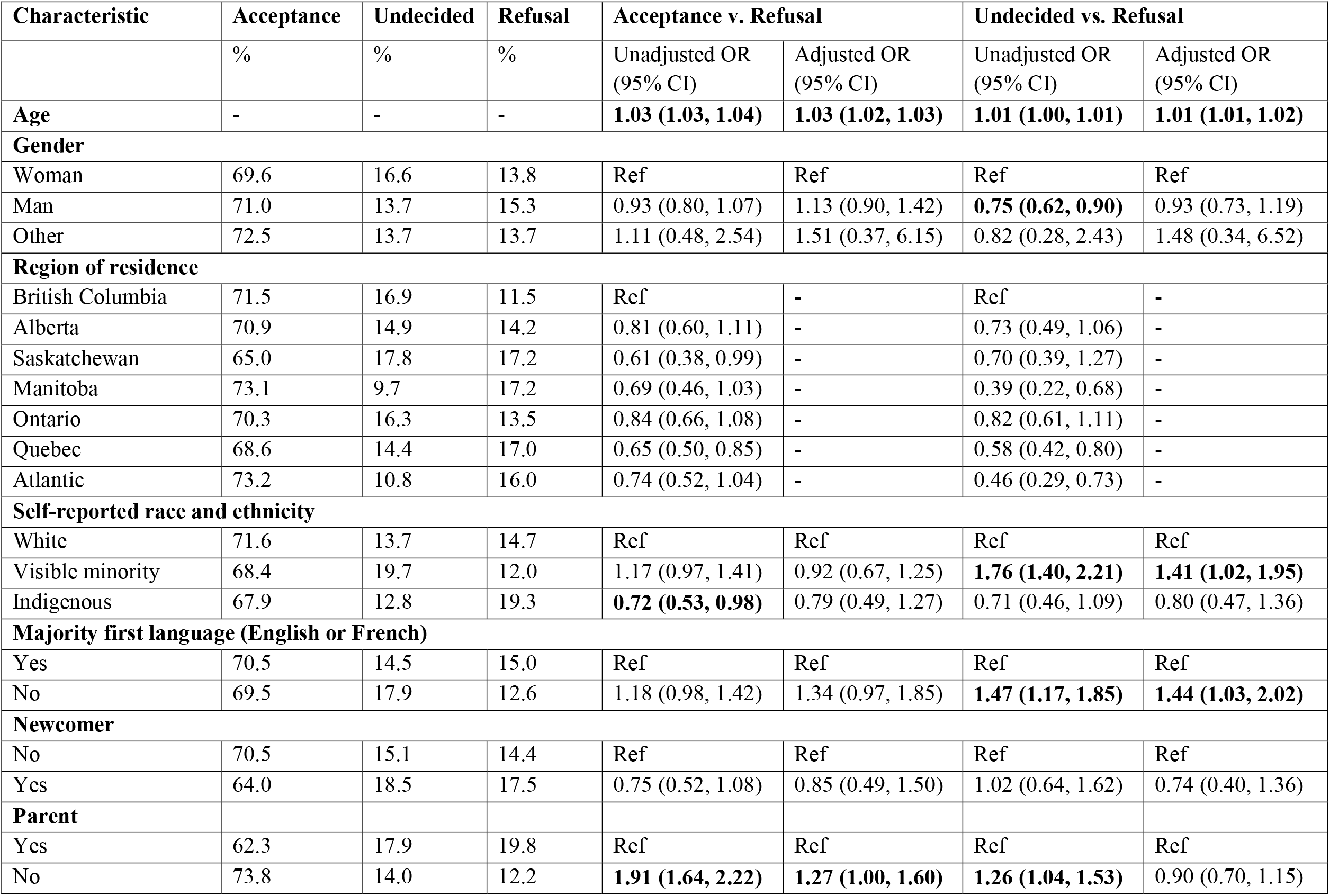
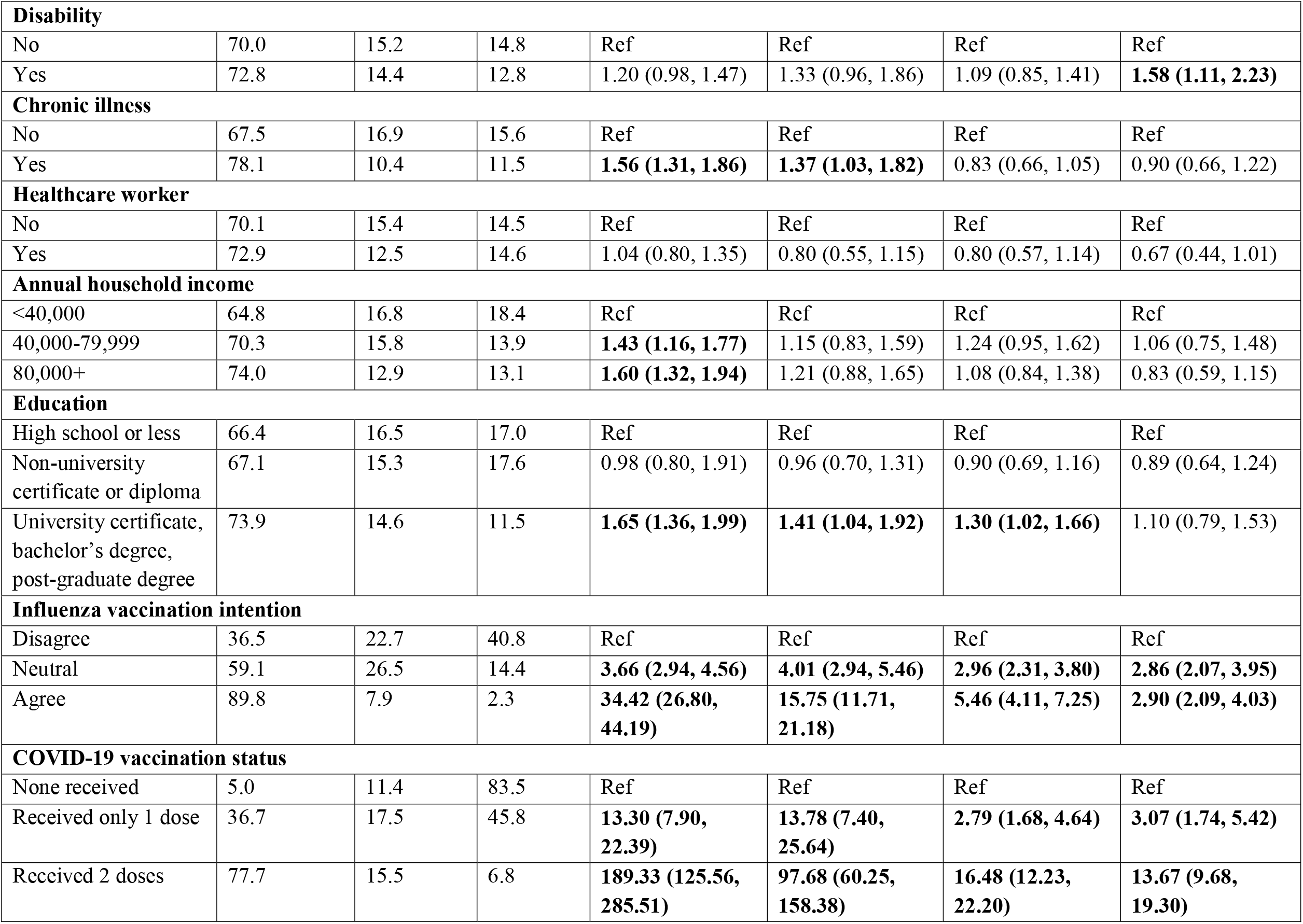
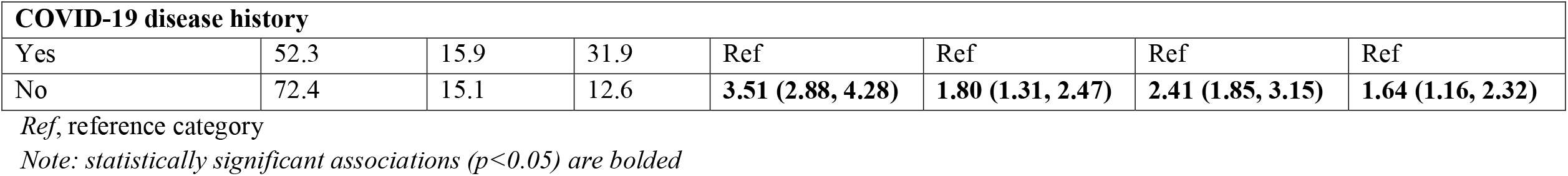
Weighted proportions and multinomial logistic regression of factors associated with COVID-19 vaccine third dose intentions

Both the acceptance and undecided groups identified government recommendations (65.1%, 44.1%), personal and/or family health reasons (57.5%, 53.5%), and healthcare provider recommendations (53.7%, 40.1%) as the most important influences on their vaccination decision-making (Fig. 1). The top three decision influences reported by the refusal group included personal and/or family health reasons (46.0%), government recommendations (33.6%) and conversations with friends and/or family (30.8%). Social media was identified as a decision influence by 6.8% of the acceptance group, 4.5% of the undecided group, and 10.8% of the refusal group. The refusal and undecided groups were most likely to specify a single influence on vaccine decision-making (67.2% and 48.6%, respectively), while the acceptance group was most likely to report three or more influences (43.1%).

**Fig. 1.**
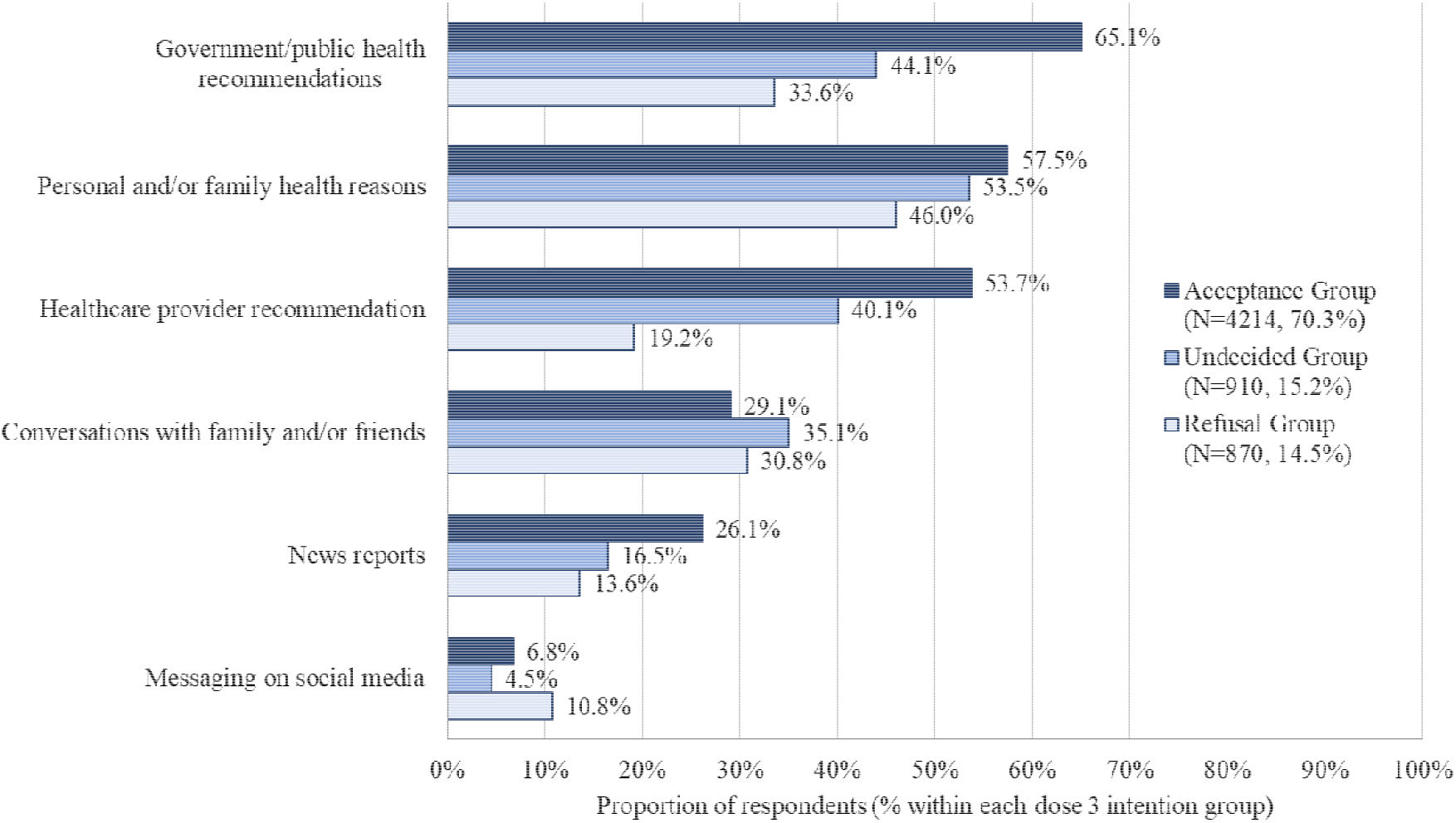
Influences on COVID-19 vaccine decision making by dose 3 intention group. Respondents were asked to select all answers that apply.

### Factors associated with accepting or being undecided about annual COVID-19 vaccination

In the multivariate model (Table 3), increasing age, minority first language, positive or neutral influenza vaccination intentions, no history of COVID-19 illness, and receipt of two doses of COVID-19 vaccine were associated with higher odds of annual dose acceptance (compared to refusal) and higher odds of being uncertain about receiving an annual dose (compared to refusal). Higher household income, higher level of education, non-parent status, presence of a chronic illness, and receipt of one COVID-19 vaccine dose were also associated with higher odds of annual dose acceptance. Presence of a disability was associated with increased odds of being undecided about annual dose receipt. Identifying with visible minority status was associated with lower odds of annual dose acceptance. When compared to female gender, male gender was associated with lower odds of being undecided about an annual dose.

**Table 3.**
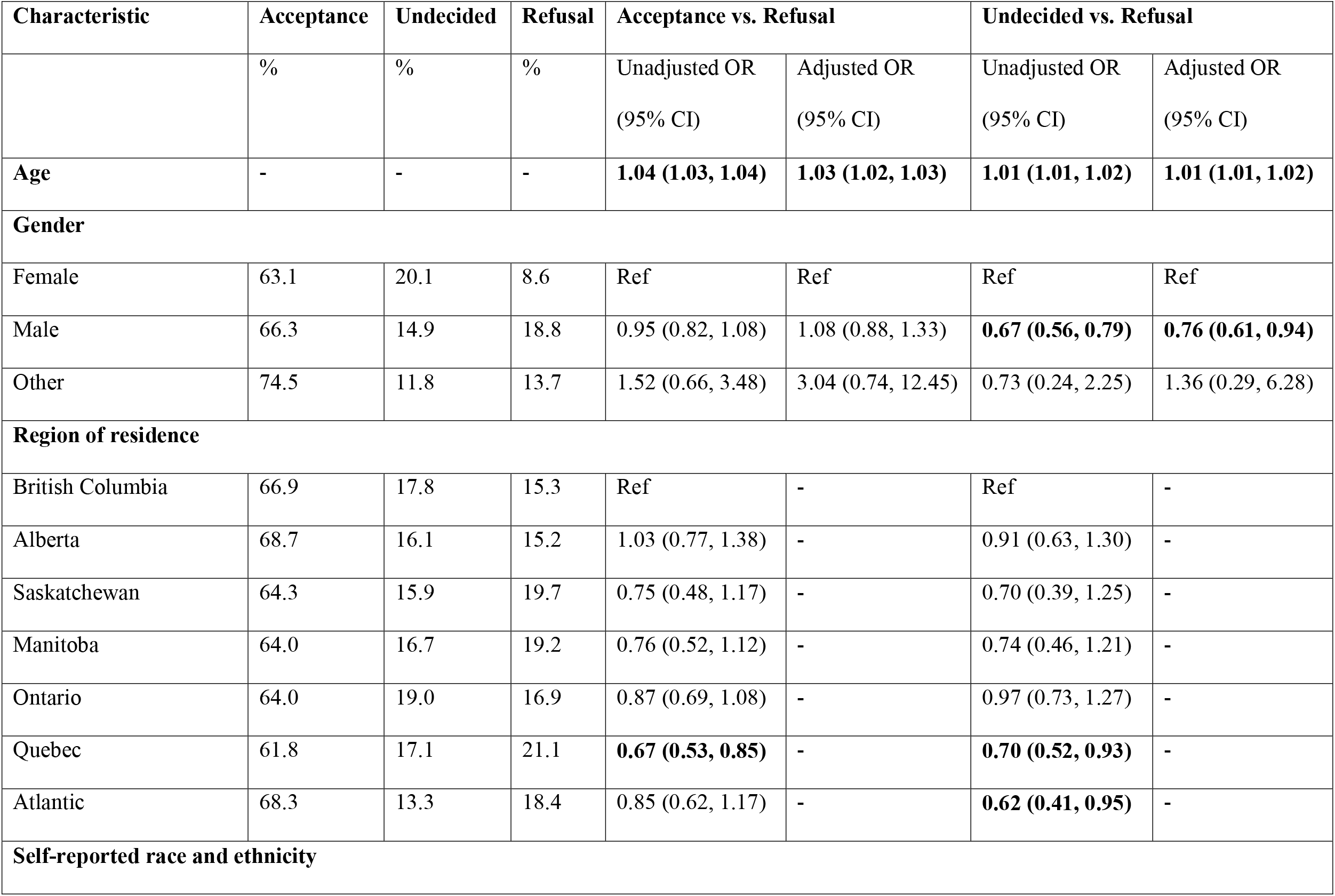
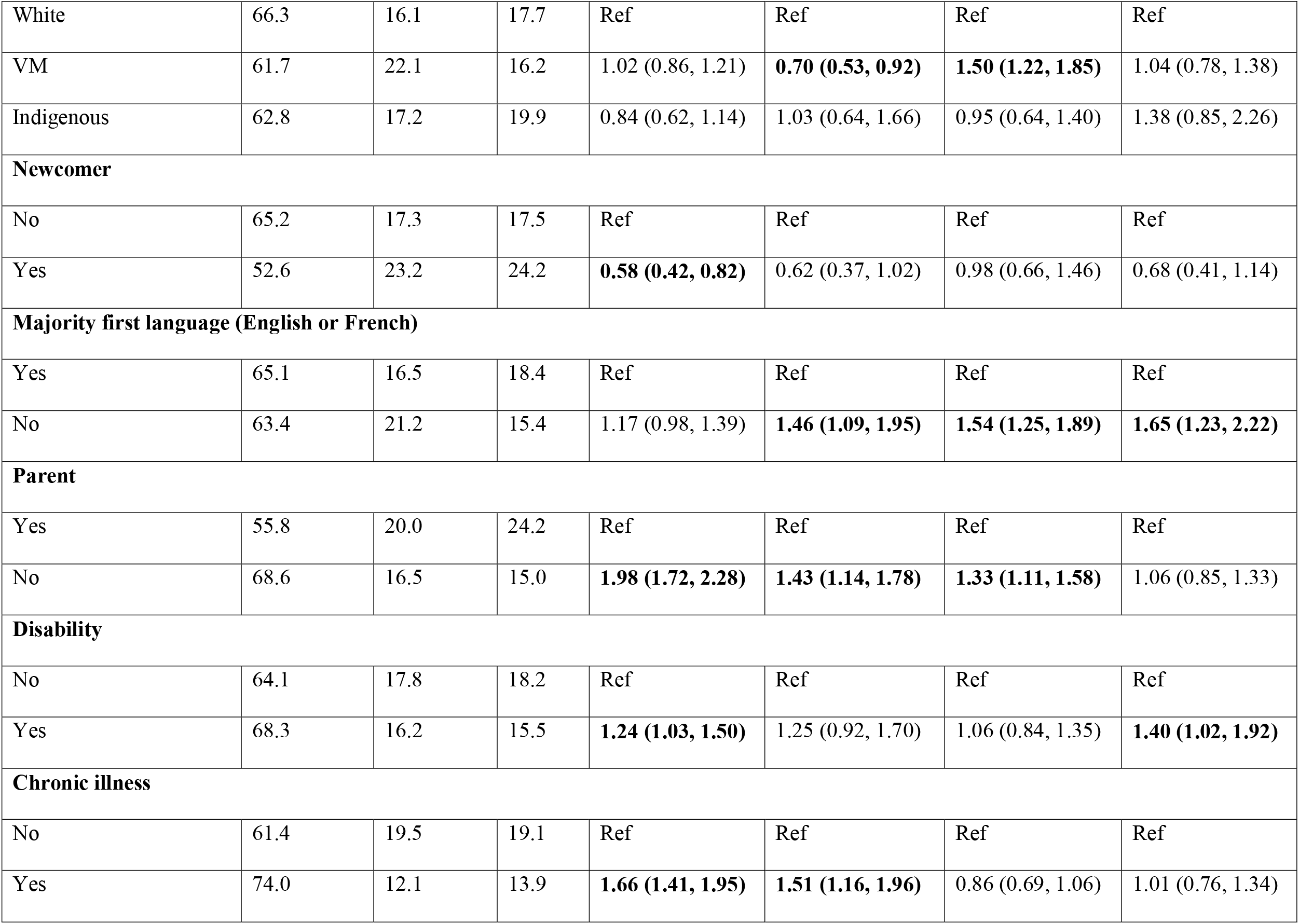
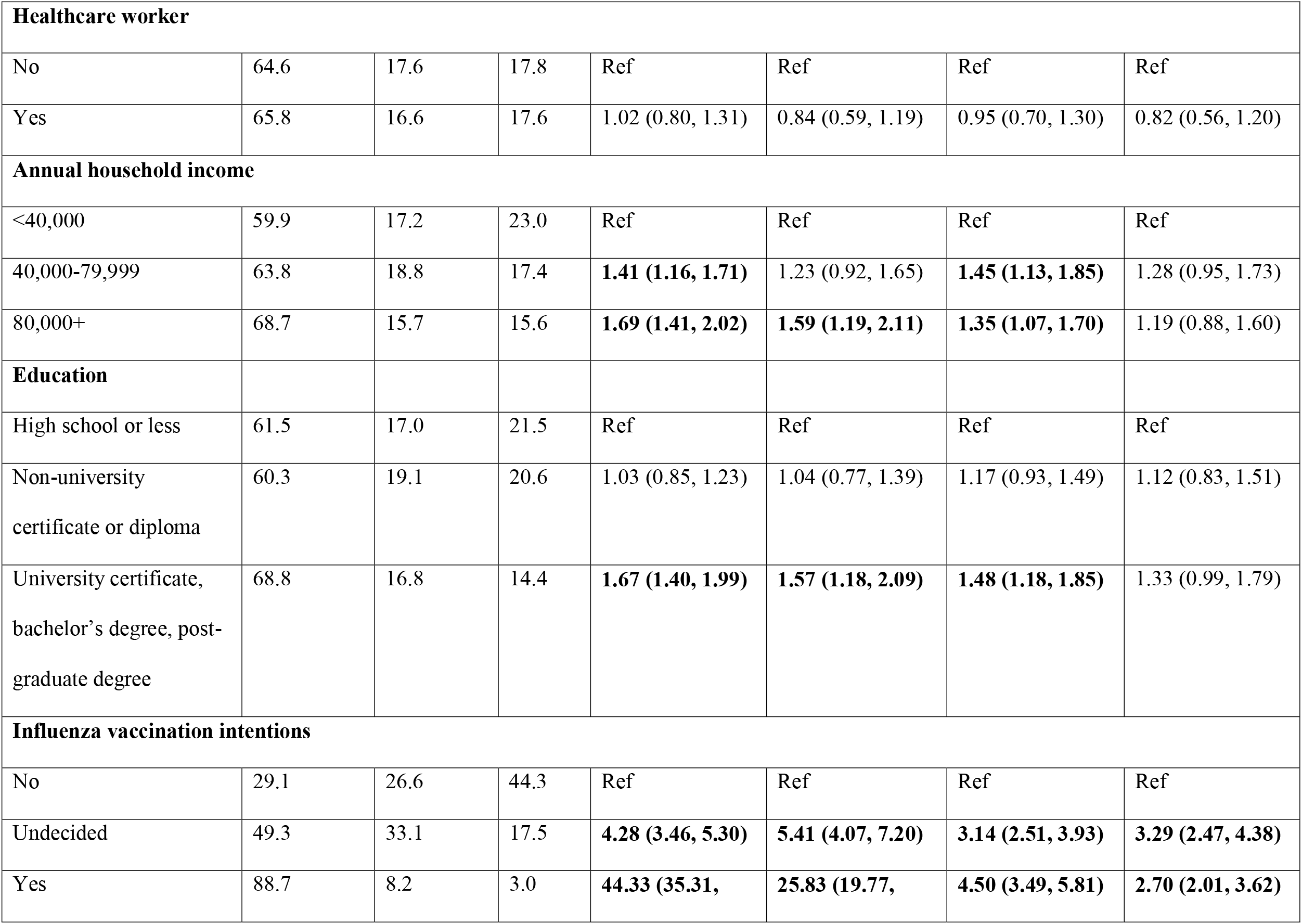
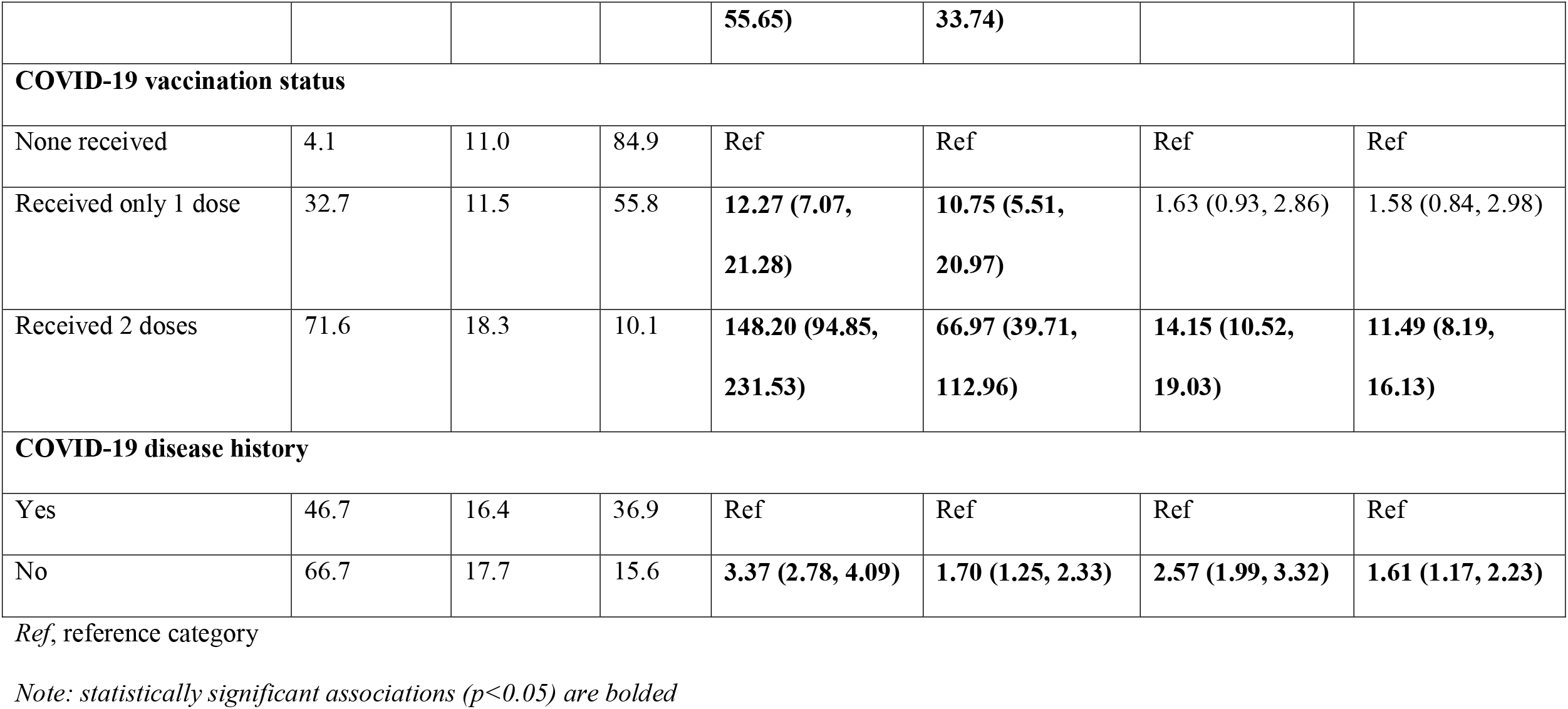
Weighted proportions and multinomial logistic regression of factors associated with COVID-19 vaccine annual dose intentions

For the annual dose intention groups, influences on vaccine decision-making were similar to patterns noted among dose 3 acceptance groups (Figure B1).

### Vaccination motivations and delivery preferences

Motivations for having previously received a COVID-19 vaccine dose for the dose 3 intention groups are presented in Figure 2. Protection of self (62.6%), protection of family (19.4%), and a desire to return to normal (6.2%) were the most commonly reported reasons for previous COVID-19 vaccine receipt among the acceptance group. Approximately 2.9% of this group identified vaccine mandates or restrictions as a main motivator. Protection of self and family were also the top two most commonly identified motivators for the undecided group (44.3% and 20.7%, respectively), with vaccine mandates or restrictions the third most commonly chosen reason (17.8%). Among the third dose rejection group, 52.3% reported they had been vaccinated because of mandates/restrictions, with 19.7% and 14.1% reporting self or family protection as the main motivator, respectively.

**Fig. 2.**
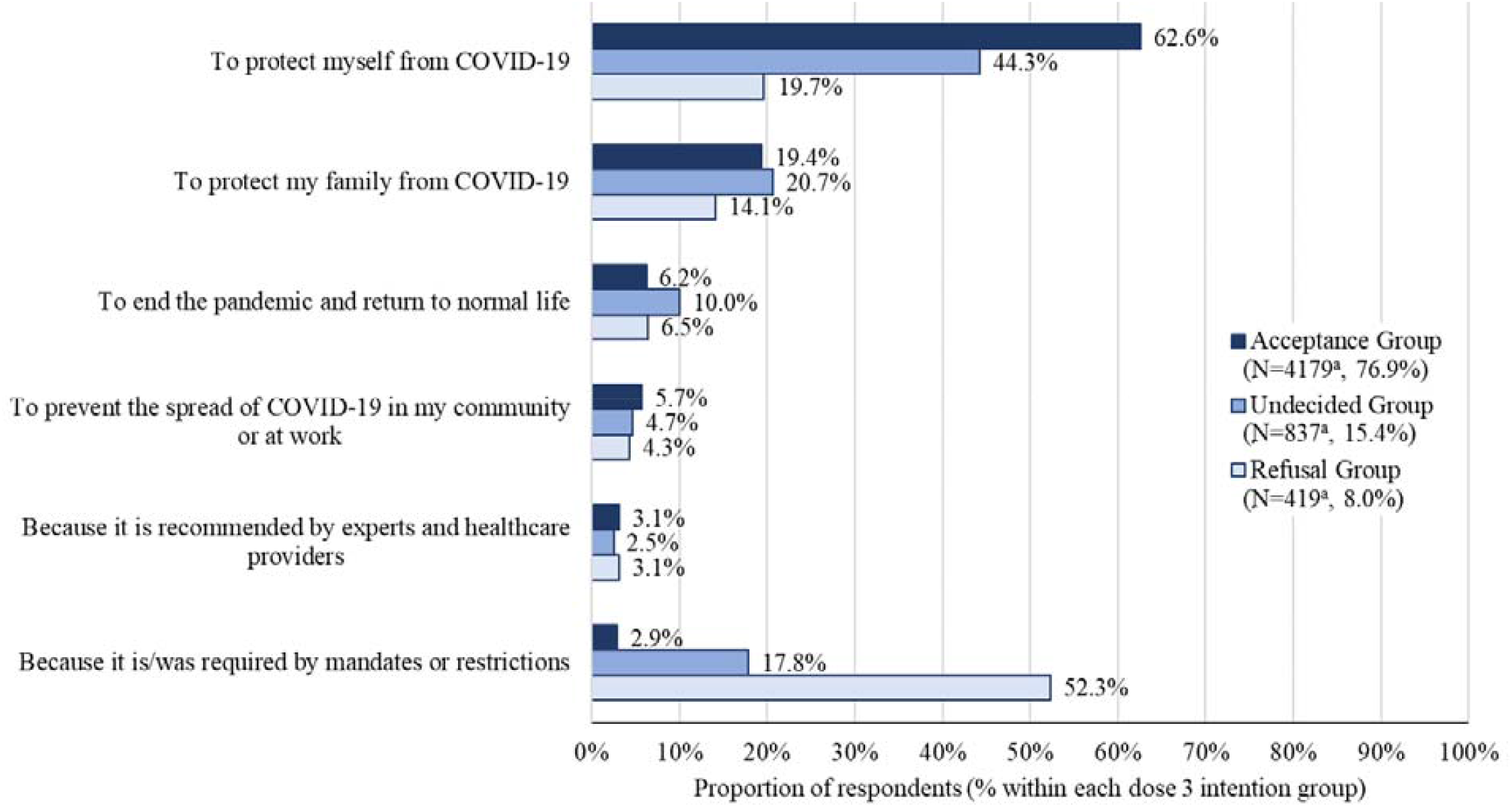
Primary motivation for previous COVID-19 vaccine receipt by dose 3 intention group. ^a^ Respondents who chose not to answer the question were removed from the denominator

The majority of the dose 3 acceptance group agreed with COVID-19 vaccine co-administration with the influenza vaccine (75.7%) or with routine vaccines (79.3%) (Fig. 3). Most of the undecided group were neutral about COVID-19 vaccine co-administration with influenza (50.7%) and routine vaccines (52.6%), while the majority of the refusal group disagreed with co-administration (with influenza (86.6%), with routine vaccines (79.5%)). Overall, 60.9% and 64.8% of respondents agreed with COVID-19 vaccine co-administration with influenza and routine vaccines, respectively.

**Fig. 3.**
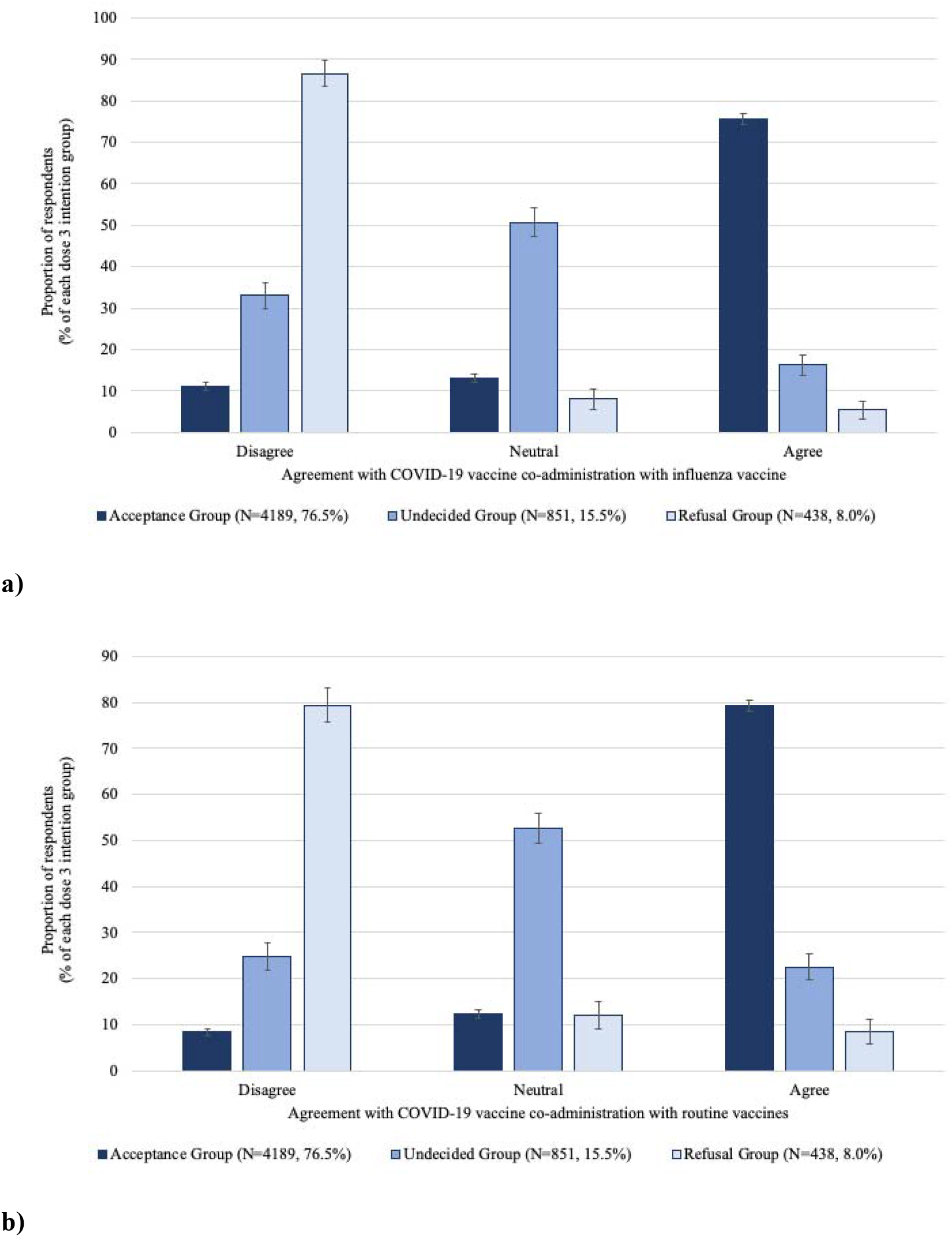
Agreement with COVID-19 vaccine co-administration with a) influenza vaccine, and b) routine vaccines, by dose 3 intention group. Included only respondents who had received at least one dose of COVID-19 vaccine.

All groups identified pharmacy as the preferred location to receive a COVID-19 vaccine (43.8% acceptance group, 40.3% undecided group, 38.6% refusal group), followed by temporary vaccination centres (23.4% acceptance group, 27.1% undecided group, 26.7% refusal group) (Fig. 4). Less than 3% of each group identified their child’s school or their home as places they would prefer to receive a vaccine.

**Fig. 4.**
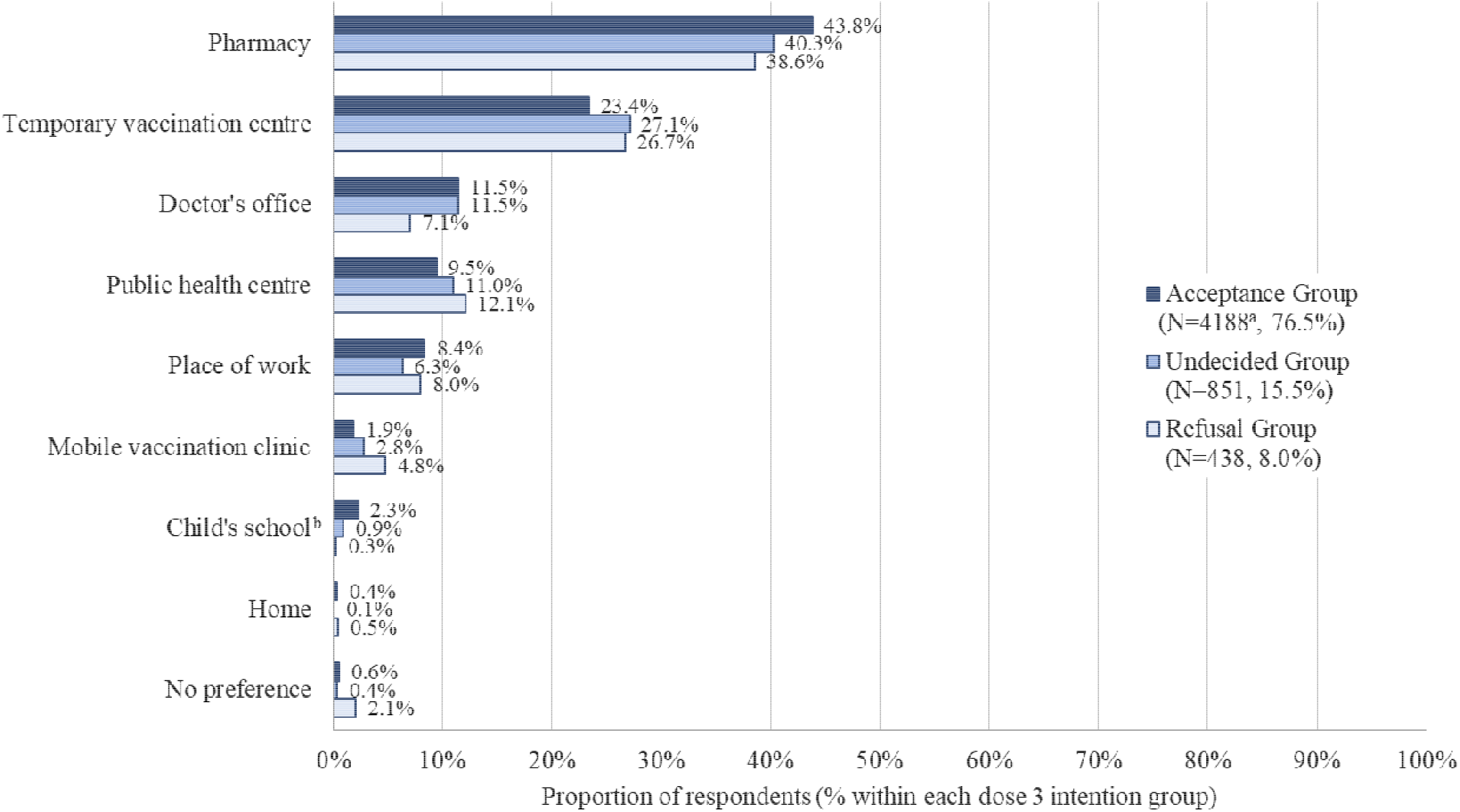
Preferred vaccination location by dose 3 intention group. Included only respondents who had received at least one dose of COVID-19 vaccine. Respondents were asked to choose one answer. ^a^ One free-text response was discarded as not relevant to the question ^b^ Proportion of parent respondents who chose this option

For all dose 3 intention groups, the most common recommendation for making vaccination easier was the ability to be vaccinated without an appointment (58.9% acceptance group, 58.4% undecided group, 55.0% refusal group), followed by close proximity to vaccination services (53.7% acceptance group, 46.3% undecided group, and 33.6% refusal group) (Figure 5). Providing childcare or allowing for family appointments was the third most commonly chosen recommendation for the acceptance and undecided groups (39.7% and 33.6%, respectively), while paid time off from work was the third most common recommendation from the refusal group (31.7%). Responses for annual dose intention groups showed similar patterns (see Appendix B).

**Fig. 5.**
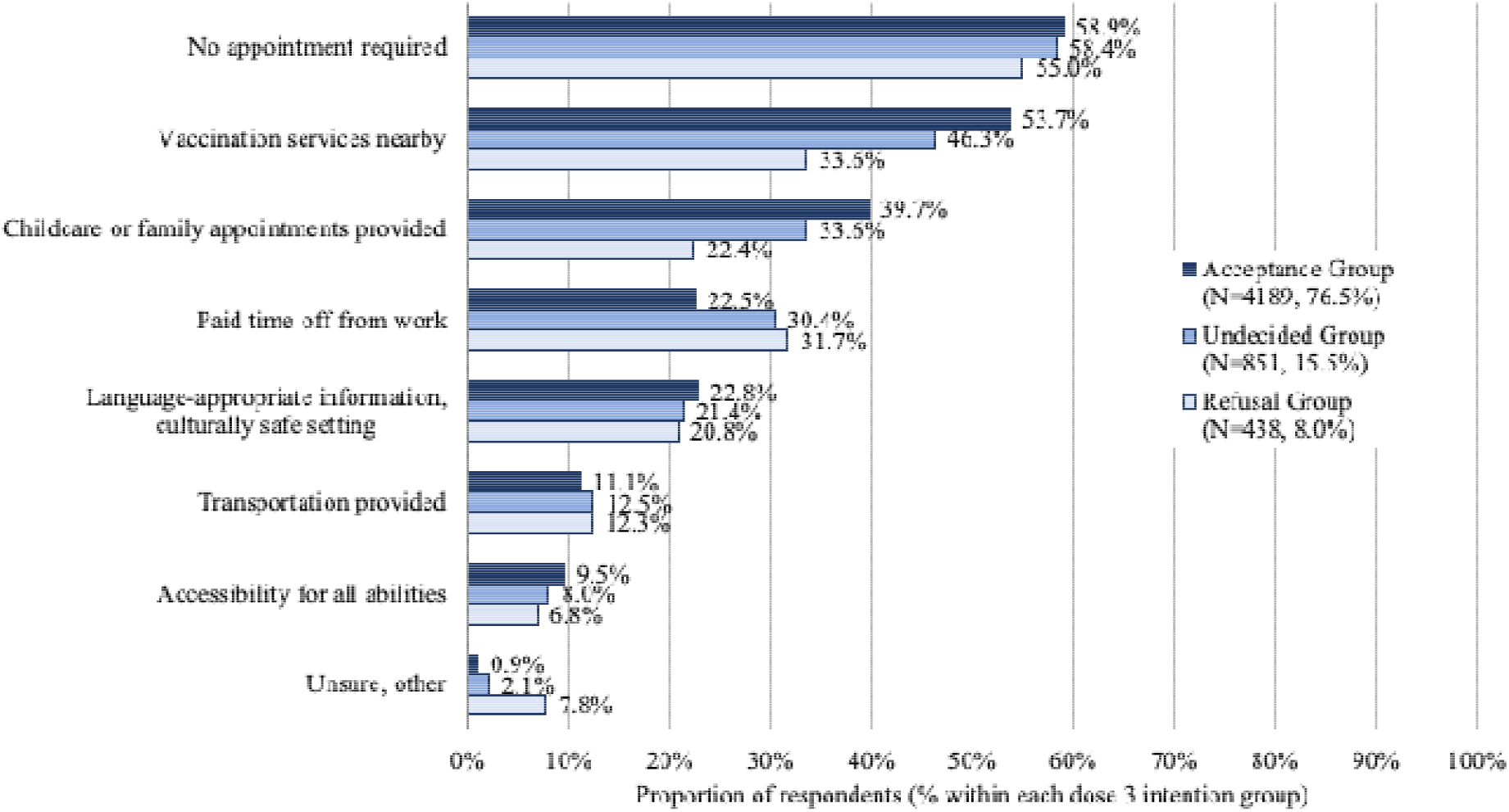
Recommendations for making the vaccination process easier by dose 3 intention group. Included only respondents who had received at least one dose of COVID-19 vaccine. Respondents were asked to select all answers that applied.

## Discussion

We completed a national, cross-sectional survey to evaluate Canadians’ intentions to receive booster (third and annual) COVID-19 vaccine doses, at a time when recommendations on additional COVID-19 vaccine doses were evolving. We found that 70% of all respondents, and 78% of those who had completed a two-dose series, indicated they would accept a third dose of COVID-19 vaccine. The proportion of two dose recipients willing to receive a third dose of vaccine was somewhat lower than findings from surveys conducted in other countries during the same time period (79-95.5%).(8-10, 21-24) However, more than 15% of respondents in our study reported they remained undecided about receiving a third dose, including over 10% of those who had not yet received any COVID-19 vaccines. Therefore, even in a population with high primary series coverage (almost 90% in our study), there is potential to improve uptake of both primary and additional doses.

Acceptance of an annual dose (65%) was lower than third dose acceptance (70%), indicating that, if COVID-19 vaccines are required on an ongoing basis, uptake may decline over time. Factors associated with accepting an annual COVID-19 vaccine were similar to third dose intentions; however, those of male gender, lower income, and visible minority identity appeared less likely to accept annual vaccination. Given that almost 20% of respondents remain undecided about annual COVID-19 vaccination, and the important influence of government and healthcare provider recommendations on vaccination decisions, acceptance is likely to be improved by clear guidelines around annual dose vaccinations, if or when they are required.

We found that many respondents with established risk factors for COVID-19 morbidity and mortality (e.g., higher age, pre-existing chronic conditions) were significantly more likely to report acceptance of additional COVID-19 vaccine doses. The relationship between increasing age and additional dose acceptance is similar to findings on third dose acceptance from other countries.(9-11, 22, 24) Literature indicates mixed results on the impact of a pre-existing chronic condition on uptake of additional COVID-19 vaccines (8-11, 23), though we defined chronic conditions more narrowly, as those at highest risk for COVID-19 morbidity/mortality. People who live with disability may also be at increased risk for both infection and negative disease outcomes, though risk varies with both type and severity of impairment (25, 26). However, self-reported disability was not associated with higher odds of additional dose acceptance in our study; instead, these respondents were more likely to be undecided about additional dose acceptance. For some respondents, this could be related to concerns around vaccine service accessibility; approximately 8% of survey respondents indicated that improved vaccine service accessibility for all abilities would be important for making vaccination easier. To ensure those who plan to receive additional vaccine doses are able to act on their intentions, and facilitate uptake in those who are undecided, vaccine services must consider the needs of all abilities(27), and involve those who live with disabilities in service planning.(28) Improved understanding of potential barriers to vaccine uptake in this population is also required.(29)

Our results indicated both concerning and encouraging relationships between sociodemographic factors and additional vaccine dose acceptance. Concerningly, lower socioeconomic status (i.e., lower household income and educational attainment) and visible minority status were related to less positive intentions toward receiving additional COVID-19 vaccines, though results were not always statistically significant. These findings are consistent with third dose vaccination intentions in the United Kingdom (10), though other research suggests socioeconomic status has little impact.(11,30) Encouragingly, other populations thought to be at risk for undervaccination (e.g. newcomers, first language not English or French, Indigenous identity) in Canada (31), reported third dose vaccination intentions that were not significantly different from the general population or more positive. Results may reflect the success of specialized public health measures within these communities, minimizing previously existing health disparities.(24) However, some inequities remain, requiring further investigations.

It is also concerning that neither parental status nor employment as a healthcare worker was associated with more positive intentions around additional COVID-19 vaccine doses, as both populations have significant potential to influence the health and vaccination status of others. Parental intention for self-vaccination is a significant predictor for COVID-19 vaccination intent for their children.(20) Healthcare workers are consistently identified as important influences on vaccine uptake decisions, both here and in other literature (32), and those who are personally vaccinated are more likely to recommend vaccination to patients.(33) However, it is important to acknowledge that there is significant heterogeneity around vaccine acceptance within both parents (20) and healthcare workers (34), though stressing altruistic reasons for receiving additional doses (e.g. protecting children, protecting patients) may be effective for both. (20, 35)

Unsurprisingly, both previous COVID-19 vaccination and seasonal influenza vaccination intention were significant predictors of favorable intentions towards additional doses. However, previous COVID-19 vaccine receipt did not uniformly predict acceptance of additional doses; more than 20% of those who had received two doses, and 60% of those who had received only one dose, did not accept additional doses. Our results indicate that COVID-19 disease history, and motivation for receiving initial doses, may explain some of that difference. In our study, those who had experienced COVID-19 disease were more likely to refuse additional vaccine doses. This relationship between previous infection and future dose refusal is not consistently found in the literature (8, 10, 11, 36), though timing of infection relative to vaccination may be important.(10,11) Given that the evidence around vaccination after SARS-CoV-2 infection is still emerging(6), confusion over the need for, and timing of, vaccination post-infection is likely. As the number of people who have experienced SARS-CoV-2 infection continues to grow, clear messaging around the effectiveness of additional vaccine doses will be required. We also found that vaccination mandates or restrictions were the main motivation for previous COVID-19 vaccine receipt for significant portions of both the refusal (almost 50% of 419 respondents) and undecided (almost 20% of 837 respondents) groups, compared to less than 3% of the acceptance group (4179 respondents). Thus, while coercive measures may positively influence initial vaccine receipt, vaccination experience is not enough to overcome hesitancy toward additional vaccine doses. This supports observations that initial hesitancy about COVID-19 vaccination appears to persist, even after two-dose completion.(10) Other factors, including belief that sufficient protection is acquired through a two-dose series (8-10), experiencing side effects from previous doses (8, 37), and concerns over receiving additional vaccine doses while other countries are struggling to secure first doses may also play a role.(38)

While our results indicate that pharmacy-based delivery and drop-in appointments may increase COVID-19 vaccine uptake, offering co-administration of the COVID-19 vaccine with influenza or routine vaccines may not have the positive impact on uptake that is expected. In general, respondent attitudes regarding vaccine co-administration mirrored their intentions for additional COVID-19 vaccine doses; however, while almost 95% of the refusal group (N=419) did not agree (disagree or neutral) with co-administration, only around three quarters of the acceptance group (N= 4179), and less than 17% of the undecided group (N=837), agreed with co-administration. Thus, while more than 60% of all respondents agreed with COVID-19 vaccine co-administration, hesitancy towards co-administration is greater than hesitancy towards the COVID-19 vaccine alone. While there is some evidence to support this finding (12), one study found that a combination influenza/COVID-19 vaccine had higher acceptance than a COVID-19 vaccine alone.(39) More work is required to understand whether and how acceptance of co-administration differs from acceptance of combination vaccines, and whether these options differentially impact uptake across populations. Providing public choice around vaccine co-administration will be important to avoid negatively impacting future uptake.

### Strengths and Limitations

Our study benefits from a large sample size that was representative of the Canadian population in age, sex, and region of residence. Another strength of our study was the targeted effort to include respondents of populations typically underrepresented in research and of particular focus for COVID-19 vaccination programs. However, there are a number of limitations associated with our sample that limit generalizability. First, our sample was drawn from a pre-existing panel with internet access who could communicate in English or French. Second, the cross-sectional design of our study prevents identification of any trends in additional vaccine dose acceptance. A number of changes to additional vaccine dose recommendations have been made in Canada since the time of our data collection, which may impact COVID-19 vaccination acceptance.

## Conclusions

Overall, intent to accept additional COVID-19 vaccine doses was high. We found that acceptance of annual COVID-19 vaccine doses was lower than acceptance for third doses, though clear guidelines and health care worker recommendations around the need for annual doses may increase uptake. Visible minority and minority language populations, and those with a disability, may be particularly receptive to interventions promoting and facilitating vaccine uptake. Efforts to promote vaccine uptake among parents of minor children and healthcare workers are needed to improve vaccine coverage overall. While approaches to facilitate access to vaccination services are important for continued uptake of COVID-19 vaccine, promoting vaccine co-administration may be less effective.

## Data Availability

Data specific to this manuscript can be requested from the senior author, provided any such requests are covered under the existing ethics approval.

## List of Abbreviations

CI: confidence interval
N/A: not applicable
Ref: reference category

## Declarations

### Ethics approval and consent to participate

The University of Alberta Health Research Ethics Board approved the study. Informed consent was obtained from all individuals who participated.

### Consent for publication

Not applicable

### Authors’ contributions

SEM, NM, MS, conceptualized the study and developed the survey; SEM, AA coordinated data acquisition; SL-P, LR conducted the statistical analysis; LR, JL, SEM interpreted data; LR, JL drafted the manuscript; SEM obtained funding and supervised the study. All authors contributed to critical revision of the manuscript and approved the final version.

## Acknowledgements

This research is part of a larger project conducted by the COVImm study team. MS is supported via salary awards from the BC Children’s Hospital Foundation and Michael Smith Health Research BC. LR is supported by a Canadian Institutes of Health Research doctoral research award.

## Appendix A

**Table A1.**
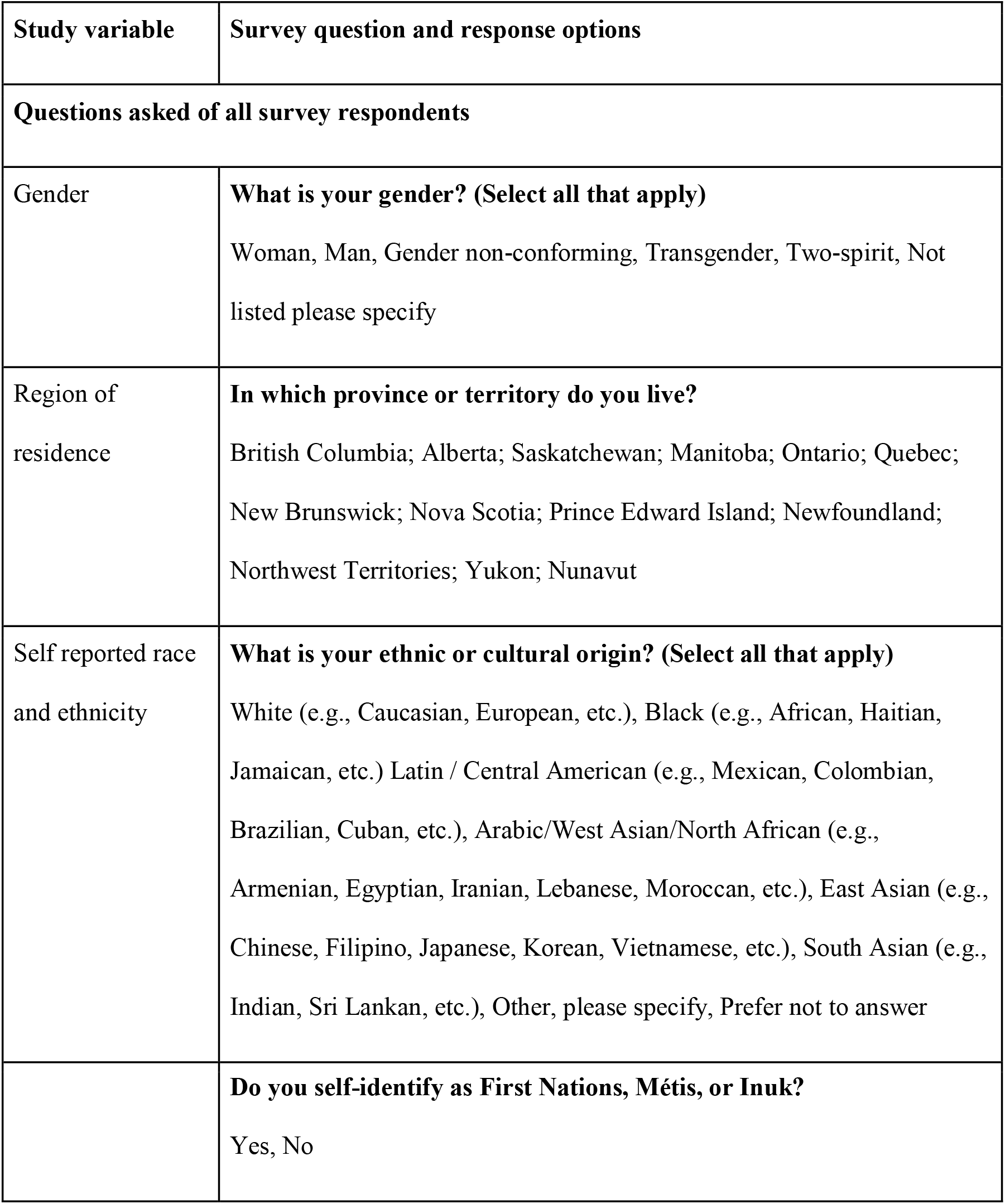
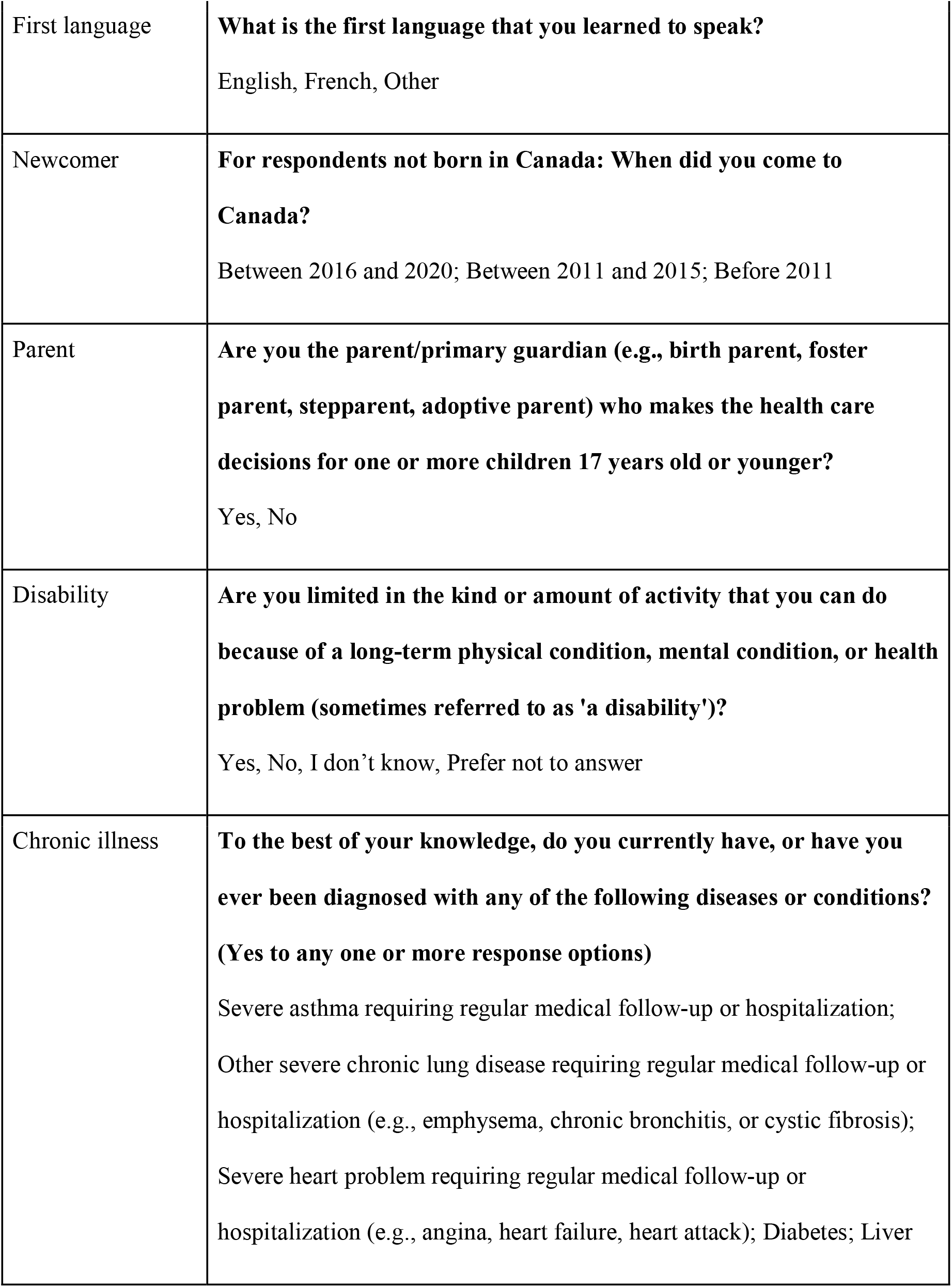
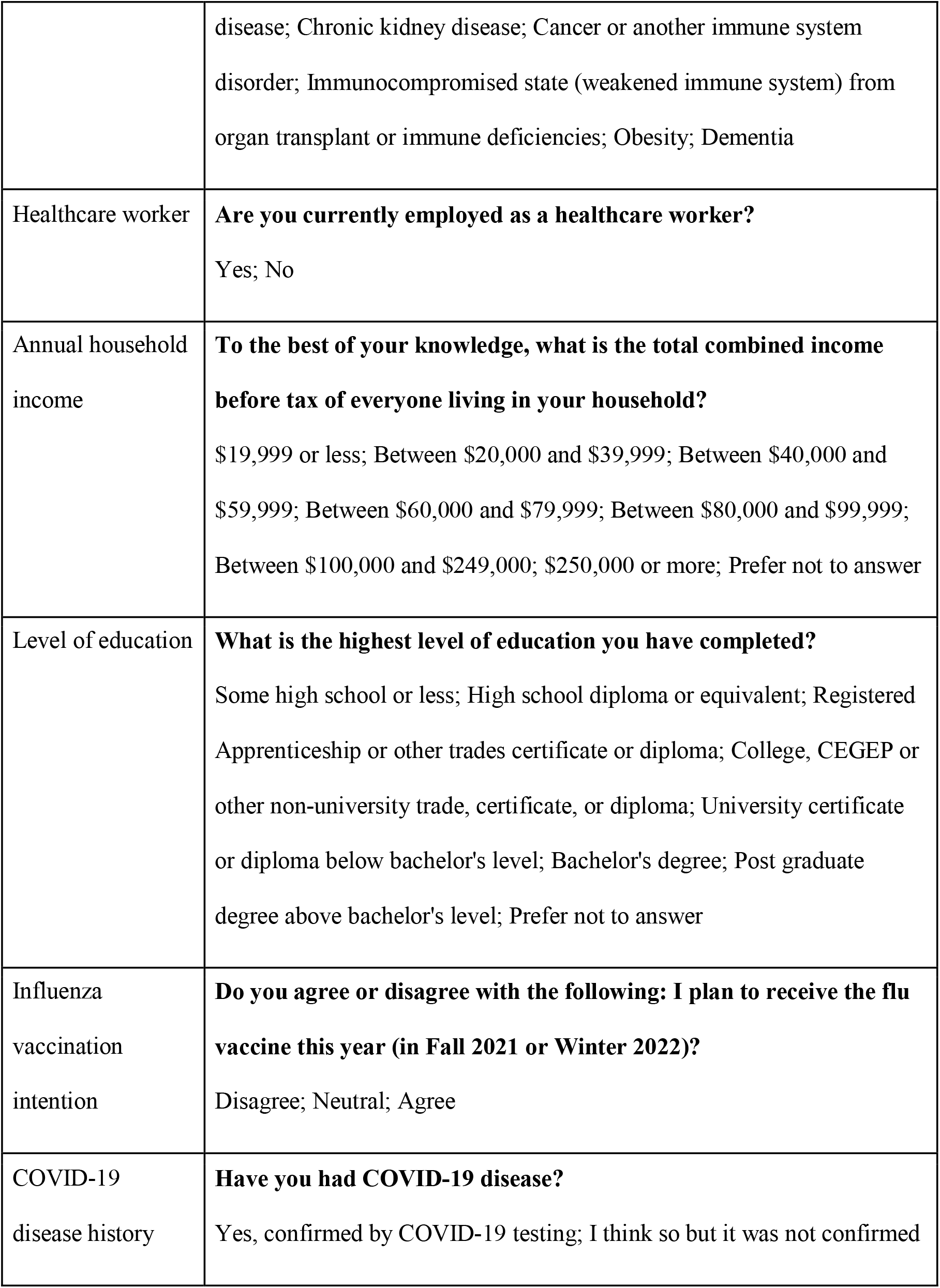
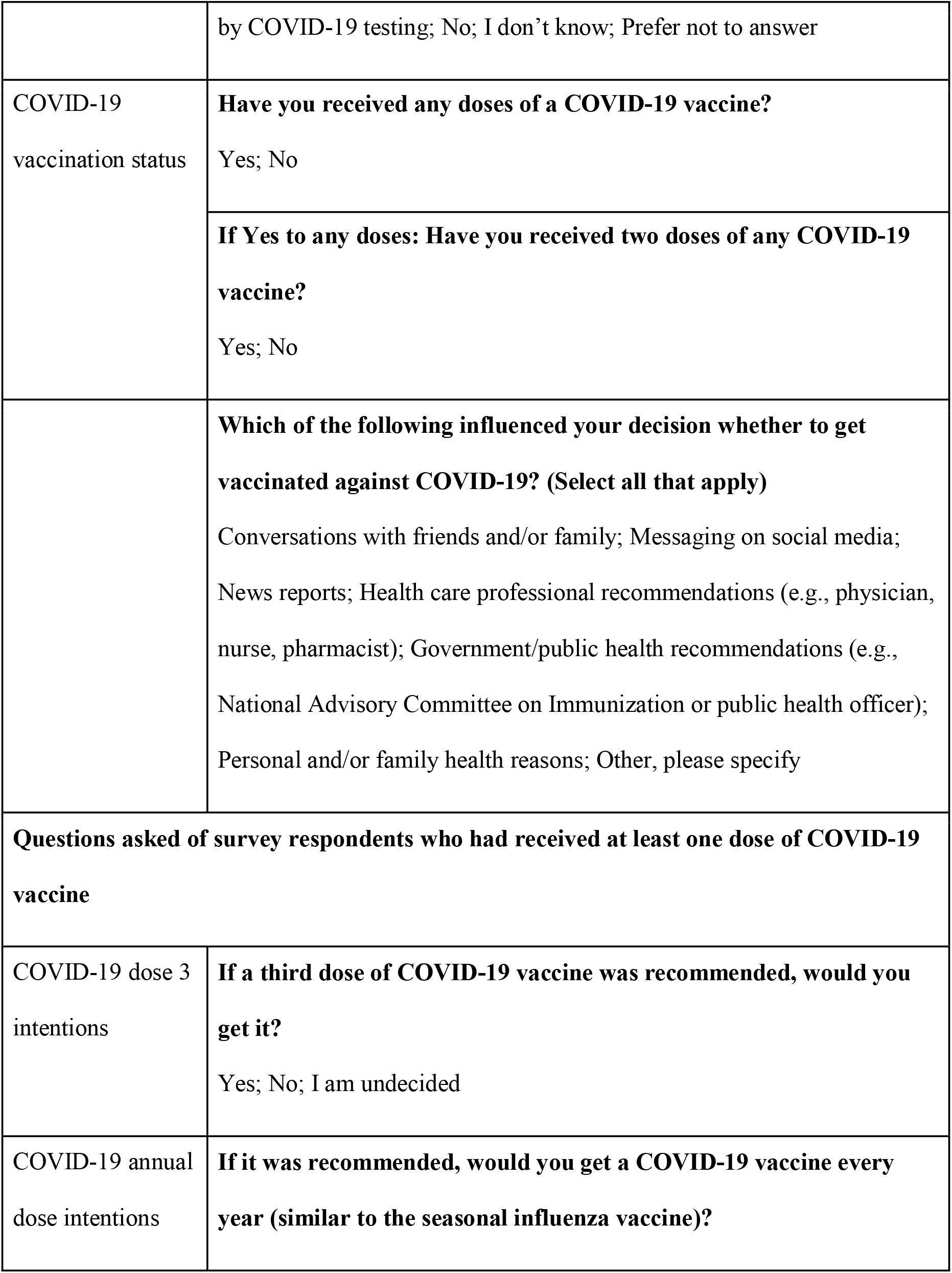
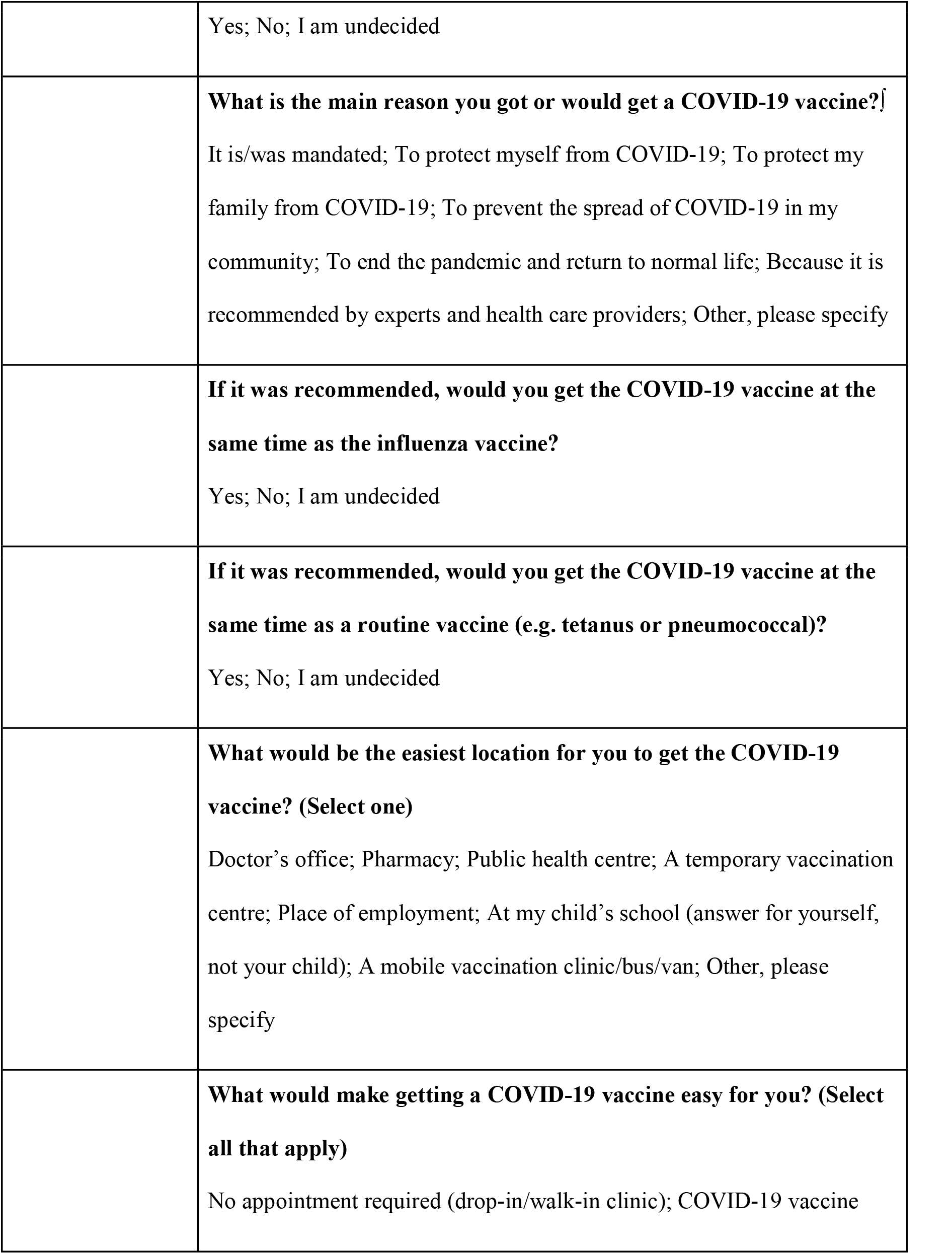
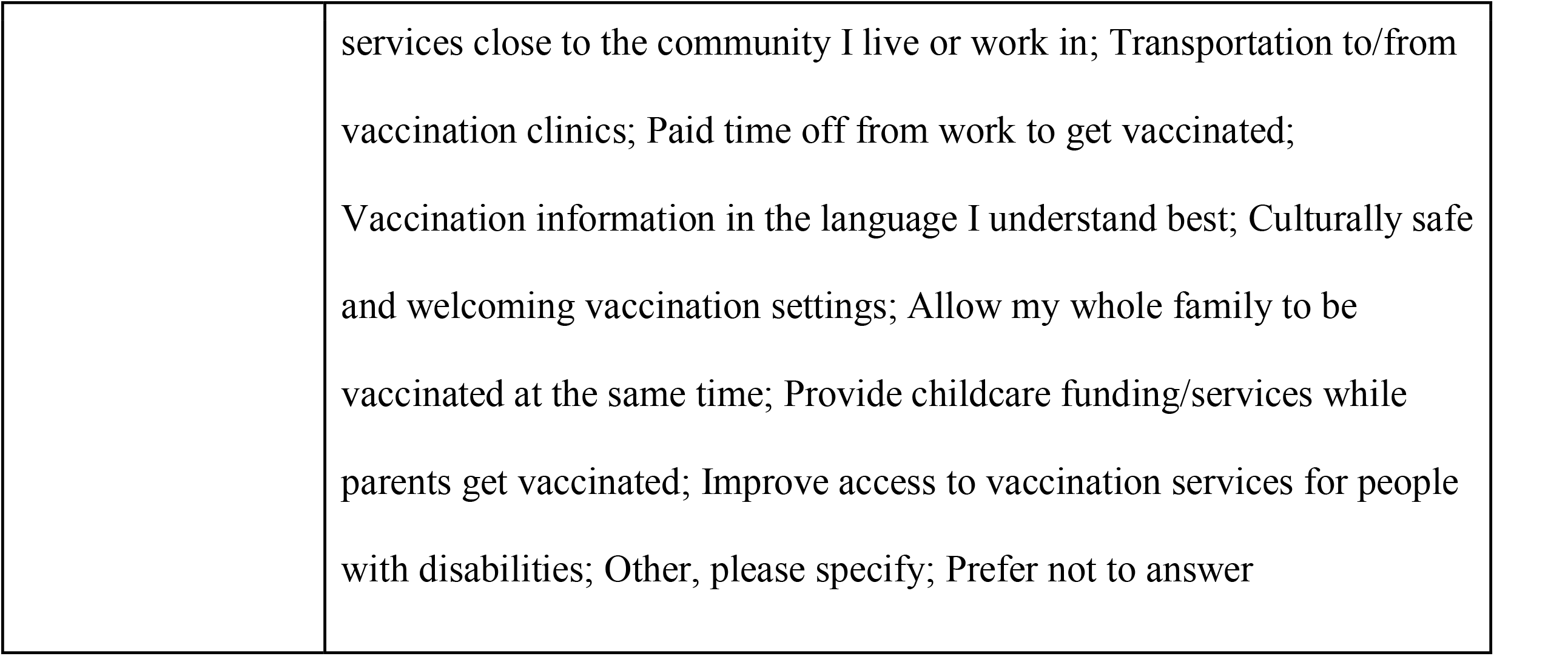
Survey questions to determine Canadian adults’ perceptions of additional COVID-19 vaccine doses and methods of delivery

## Appendix B

**Fig. B1.**
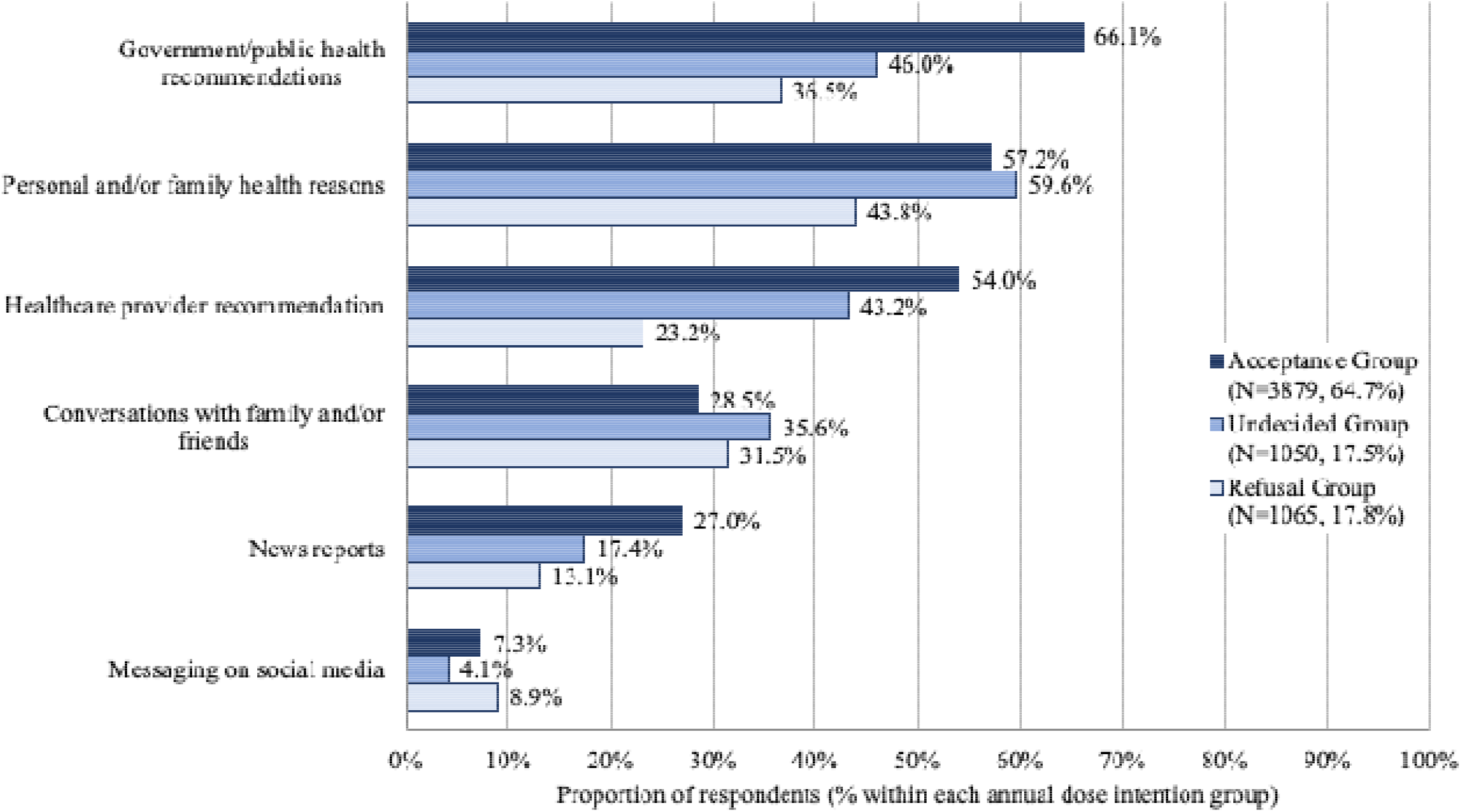
Influences on COVID-19 vaccine decision making by annual dose intention group. Respondents were asked to select all answers that apply.

**Fig. B2.**
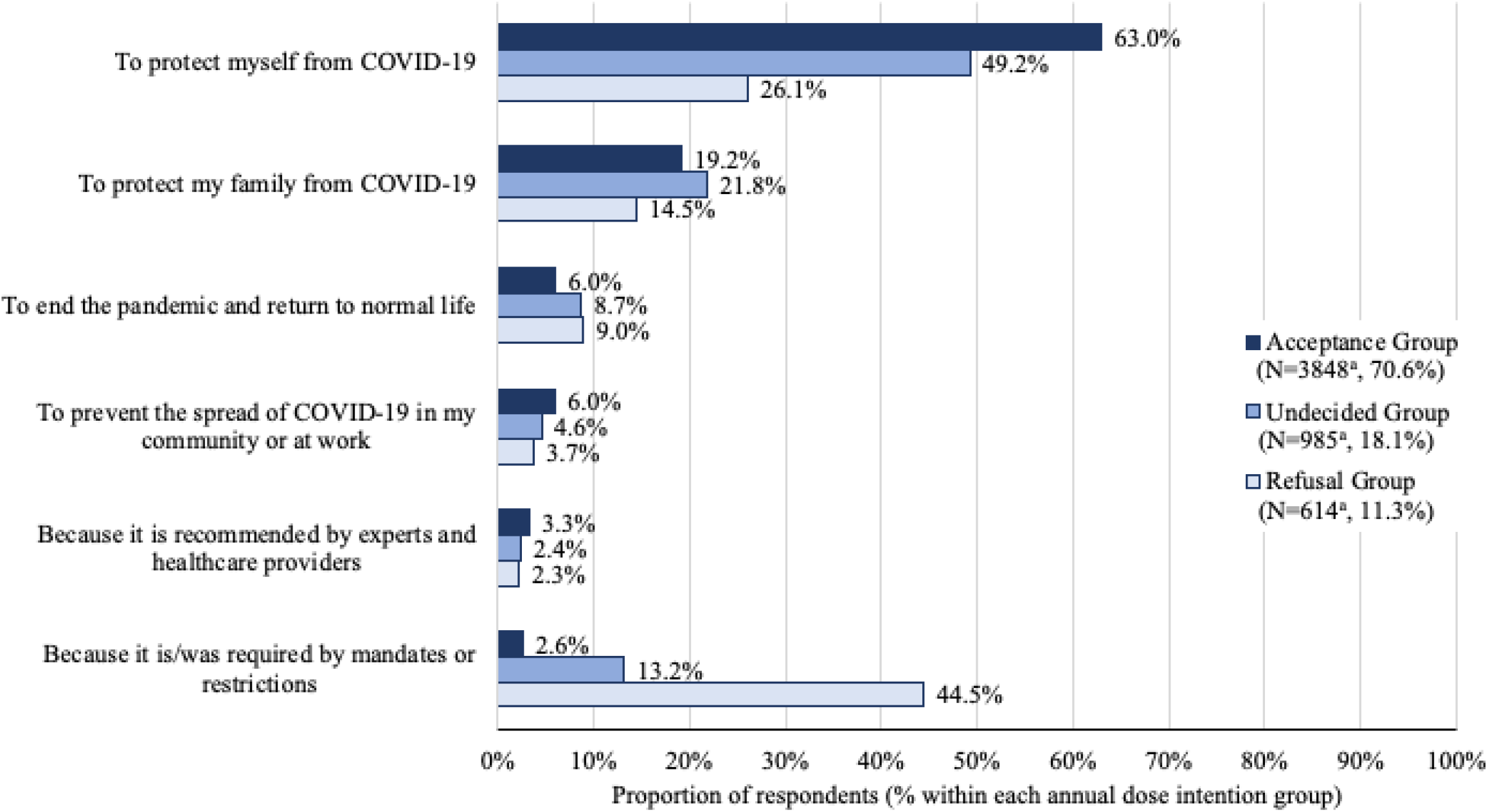
Primary reason for previous COVID-19 vaccination by annual dose acceptance group. ^a^ Respondents who chose not to answer the question were removed from the denominator

**Fig. B3.**
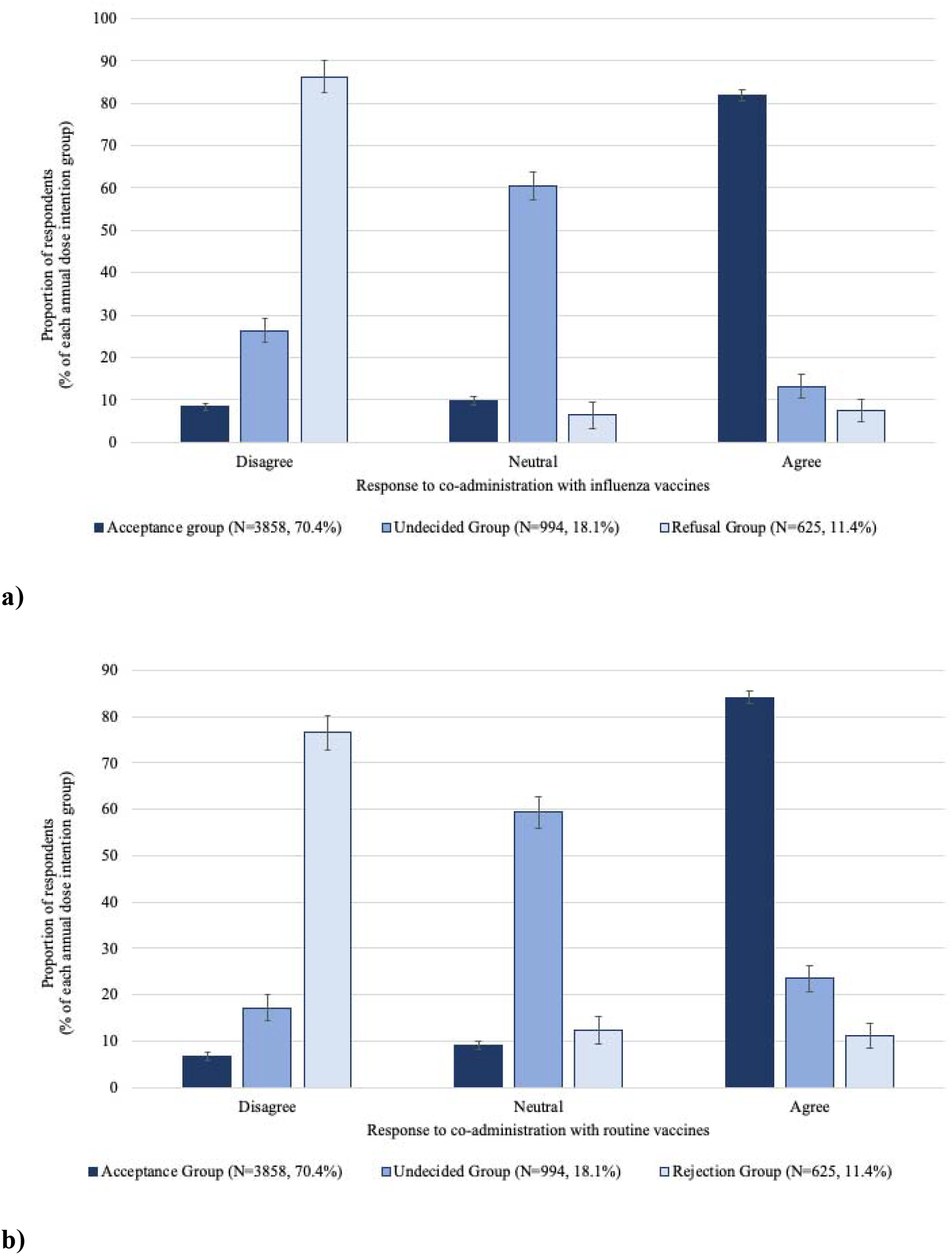
Perception of COVID-19 vaccine co-administration with a) influenza vaccine, and b) routine vaccines, by annual dose intention group. Included only respondents who had received at least one dose of COVID-19 vaccine.

**Fig. B4.**
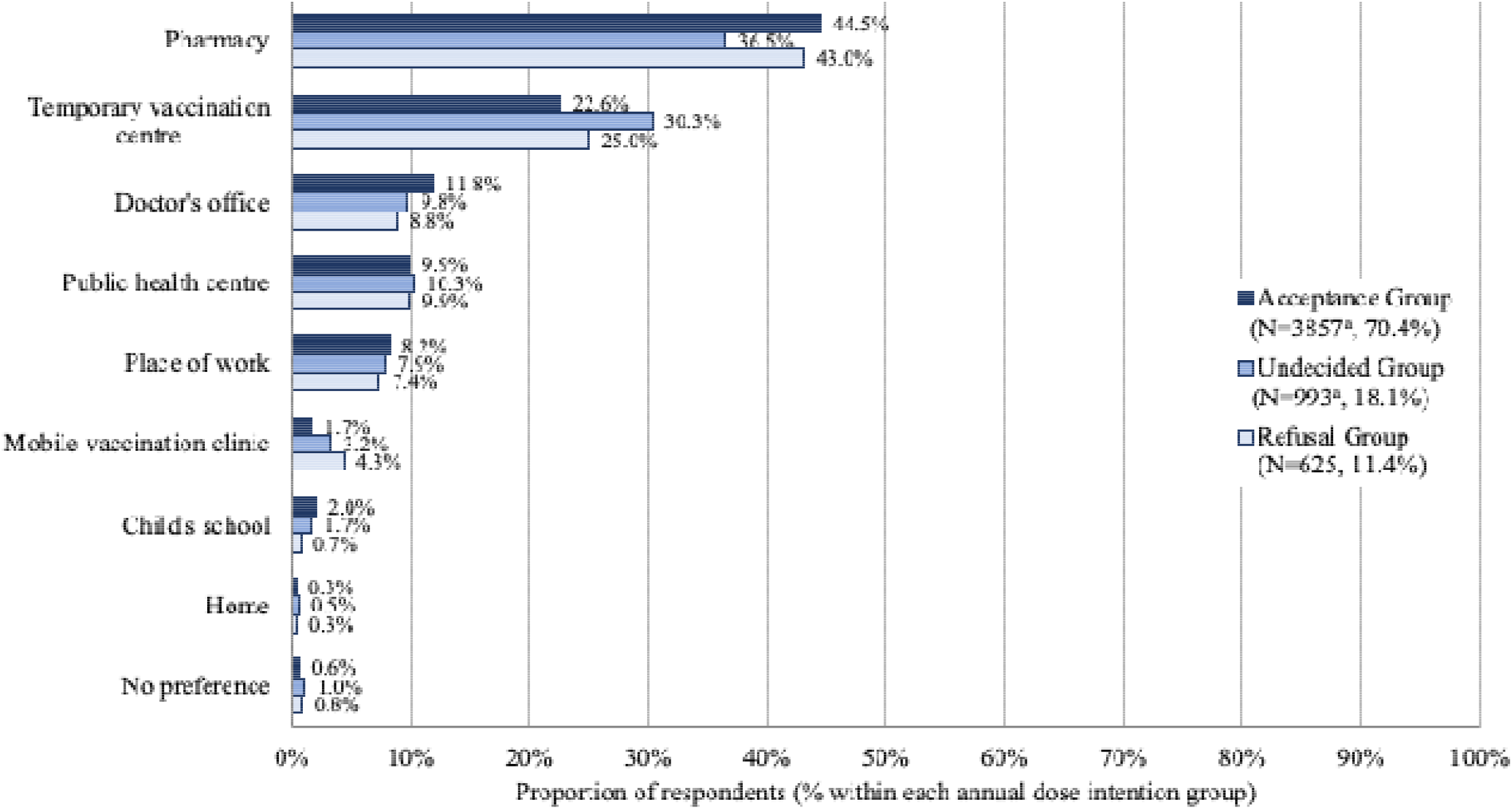
Preferred vaccination location by annual dose intention group. Included only respondents who had received at least one dose of COVID-19 vaccine. Respondents were asked to choose one answer. ^a^ One free-text response was discarded as not relevant to the question

**Fig. B5.**
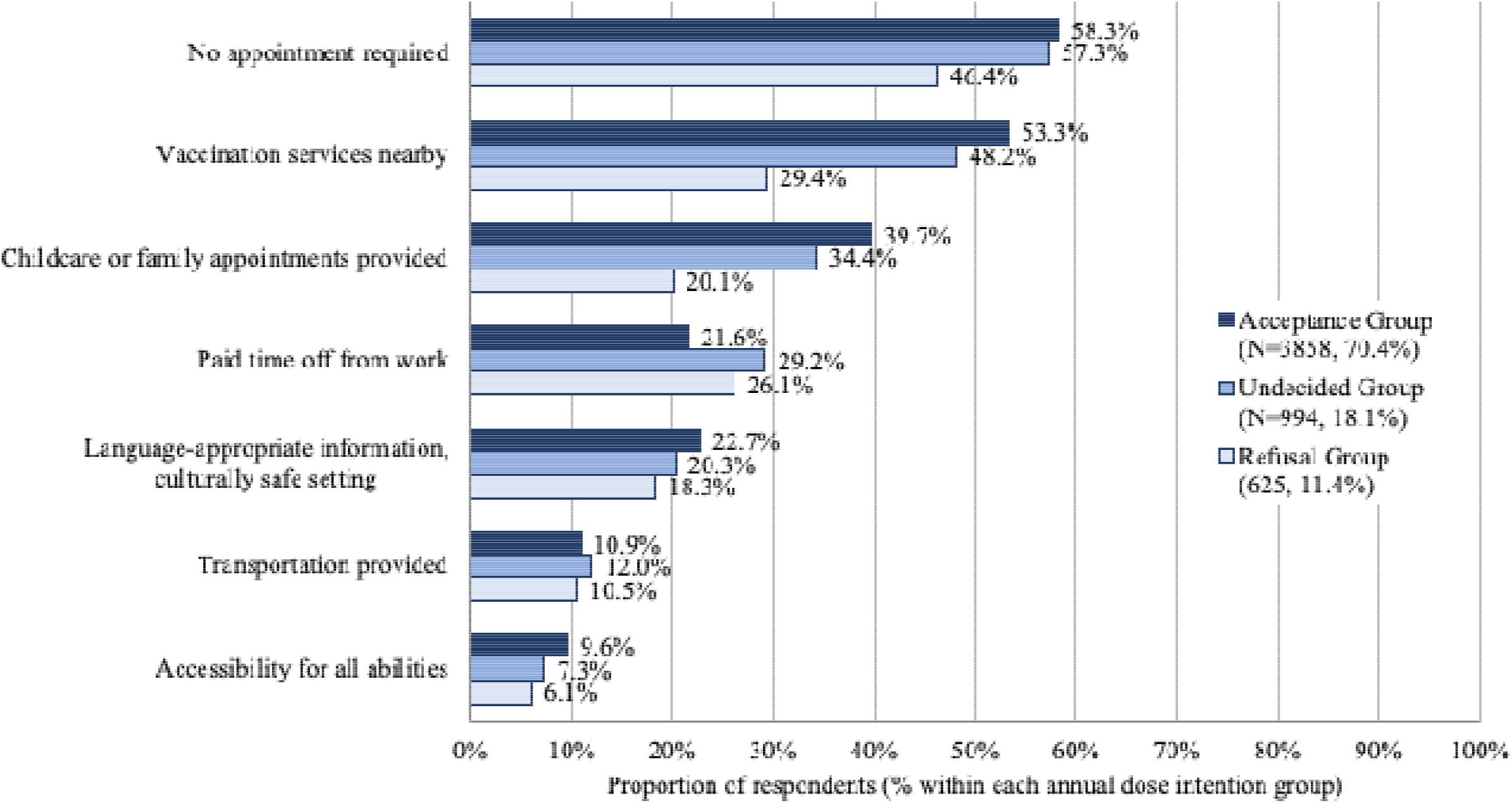
Recommendations for making the vaccination process easier by annual dose intention group. Included only respondents who had received at least one dose of COVID-19 vaccine. Respondents were asked to select all answers that applied.

